# Impact of moderate alcohol consumption on brain iron and cognition: observational and genetic analyses

**DOI:** 10.1101/2022.02.07.22270583

**Authors:** Anya Topiwala, Chaoyue Wang, Klaus P. Ebmeier, Stephen Burgess, Steven Bell, Daniel F. Levey, Hang Zhou, Celeste McCracken, Adriana Roca, Steffen E. Petersen, Betty Raman, Masud Husain, Joel Gelernter, Karla L. Miller, Stephen M. Smith, Thomas E. Nichols

## Abstract

Brain iron deposition has been linked to several neurodegenerative conditions and reported in alcohol dependence. Whether iron accumulation occurs in moderate drinkers is unknown. Our objectives were to investigate causal relationships between alcohol consumption and brain iron levels and to examine whether higher brain iron represents a potential pathway to alcohol-related cognitive deficits.

Multi-organ susceptibility-weighted magnetic resonance imaging was used to ascertain iron content of brain (quantitative susceptibility mapping (QSM) and T2*) and liver tissues (T2*), a marker of systemic iron. Observational associations between brain iron markers and self-reported alcohol consumption (n=22,254 UK Biobank participants) were compared with associations with genetically-predicted alcohol intake from two-sample Mendelian randomization (MR). Potential pathways to alcohol-related iron brain accumulation through elevated systemic iron stores (liver) were explored in causal mediation analysis. Cognition was assessed at the time of scan and in online follow-up.

Alcohol consumption was associated with markers of higher iron in putamen (β=0.09 standard deviation (S.D.) [95% confidence interval 0.07 to 0.10]), caudate (β=0.06 [0.04 to 0.07]) and substantia nigra (β=0.04 [0.02 to 0.05]), and lower iron in the thalami (β= -0.05 [-0.06 to -0.03]). Quintile-based analyses found these effects in those consuming >7 units (56g) alcohol weekly. MR analyses provided some evidence these relationships are causal. A 1 S.D. higher genetically-predicted number of alcoholic drinks weekly associated with 0.25 [0.01 to 0.49] S.D. higher putamen susceptibility and 0.28 [0.05 to 0.50] S.D. higher hippocampus susceptibility. Genetically-predicted alcohol use disorder was causally related to higher putamen susceptibility (0.18 [0.001 to 0.35] S.D.), serum iron (0.12 [0.05 to 0.19] S.D.) and transferrin saturation (0.11[0.03 to 0.19] S.D.). Elevated liver iron was observed at just >11 units (88g) alcohol weekly (0.03 mg/g [0.01-0.05] higher c.f. <7 units (56g) weekly). Systemic iron levels partially mediated associations of alcohol intake with brain iron accumulation (proportion mediated: 32% [22 to 49]). Markers of higher basal ganglia iron were associated with accelerated age-related decline in executive function and fluid intelligence, and slower reaction times.

This study represents the largest investigation of moderate alcohol consumption and iron homeostasis to date. Even low levels of alcohol consumption were associated with iron accumulation in the brain, suggesting a potential mechanism for alcohol-related cognitive decline.

## Introduction

There is growing evidence that moderate alcohol consumption adversely impacts brain health, contradicting earlier claims [1, 2]. Given its prevalence, there are huge public health implications [3]. Clarity about the pathological mechanisms by which alcohol acts upon the brain is vital not just for disease aetiology, but also to offer opportunities for intervention.

One largely neglected possibility is that iron overload contributes to alcohol-related neurodegeneration [4, 5]. Whilst neurological sequelae of inherited iron overload disorders have long been recognised [6], higher brain iron has now been implicated in the pathophysiology of Alzheimer’s and Parkinson’s diseases [7, 8]. Intriguingly, not only does the clinical profile of alcohol-related dementia overlap with these disorders, but recent observational evidence suggests heavy alcohol use may associate with iron accumulation in the brain [9, 10].

What has not previously been explored is whether brain iron accumulation is observed with moderate alcohol consumption, and if so, whether these associations are causal. Furthermore, the mechanisms by which alcohol could influence brain iron, and whether there are clinical consequences of subtle elevations in brain iron are unknown. Low levels of drinking have been observationally associated with blood markers of iron homeostasis. However, studies to date have been small, have neglected genetic contributions to iron accumulation (polymorphisms predicting serum iron are highly prevalent in European populations [11]), and serum markers may be poorly specific for body iron stores [12-14]. Better insights require well-powered samples with genetic data and concurrent measurement of iron accumulation in brain and liver, the most reliable indicator of the body’s iron stores [15, 16]. If iron deposition is mechanistically involved in alcohol’s effect on the brain, there are potential opportunities for earlier monitoring via serum iron markers, as well as intervention with chelating agents [17, 18].

In this study, we performed the largest multi-organ investigation into alcohol-related iron homeostasis to date. Alcohol consumption weekly was divided into quintiles, and never and previous drinkers distinguished. Conventional observational and genetic analyses were triangulated to investigate causal effects (**Figure 1**). Brain iron content was ascertained using quantitative susceptibility mapping (QSM) and T2*, both derived from susceptibility-weighted magnetic resonance imaging (swMRI) data. T2* reflects differences in tissue microstructure related to iron (sequestered to ferritin) and myelin, and correlates with post-mortem estimates of iron deposits in brain grey matter [19]. Susceptibility reflects the net (sequestered and non-sequestered) content of susceptibility-shifting sources like iron and myelin. Systemic iron levels were estimated using MRI-derived liver iron (a marker of liver iron levels [20]), in addition to genetically predicted serum markers. Our aims were to:

1. characterise the dose-response relationship of alcohol consumption) and brain iron in observational and Mendelian randomization frameworks,
2. investigate whether alcohol influences brain iron via changes in systemic iron levels,
3. examine whether higher brain iron represents a potential pathway to alcohol-related cognitive deficits.

**Figure 1:**
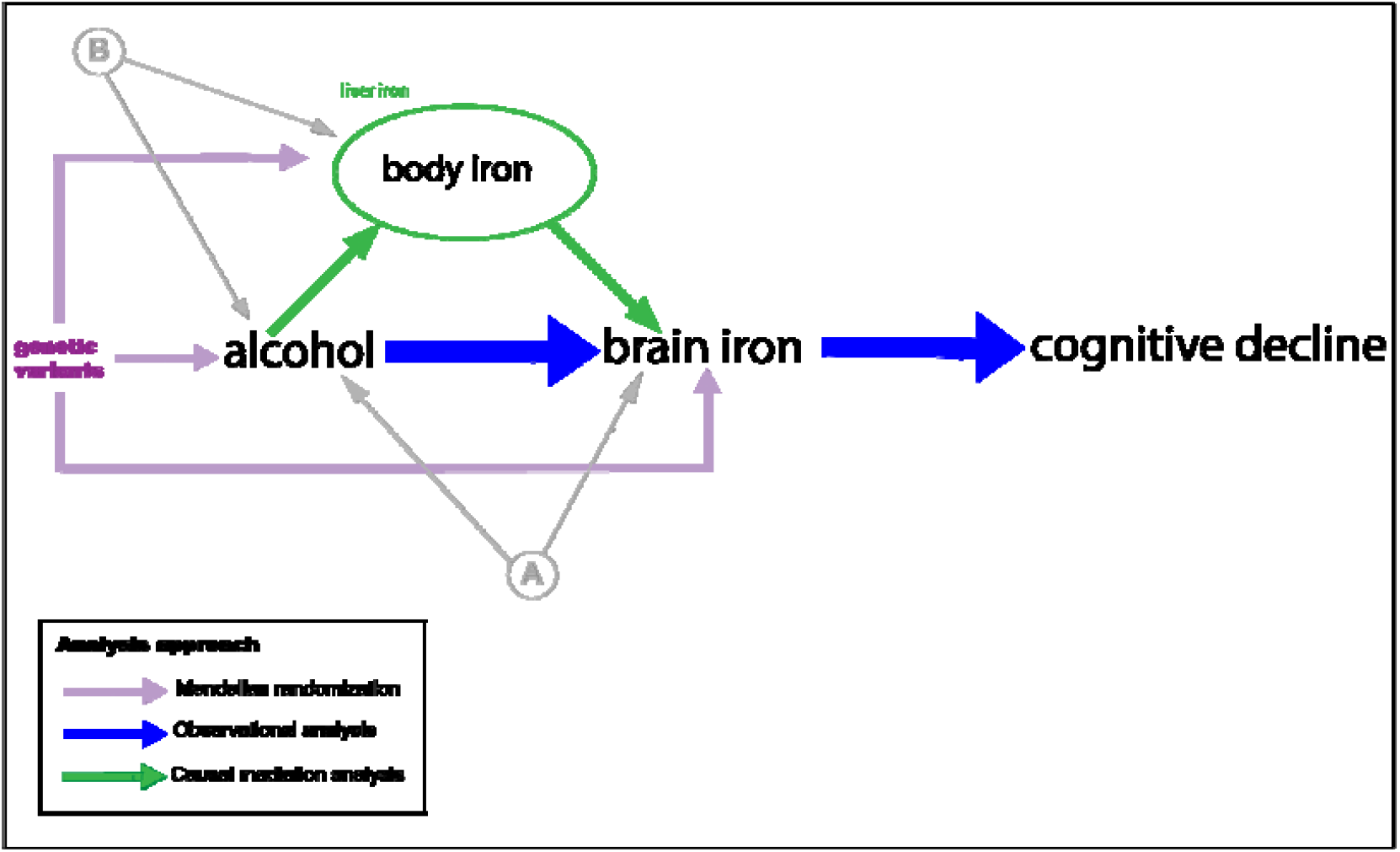
Hypothesised model and analysis approach. (A) and (B) represent unmeasured confounding. Brain iron is measured using T2* and susceptibility (UK Biobank), and body iron stores proxied using liver T2* (UK Biobank).

## Methods

### Participants

Participants were scanned as part of the UK Biobank study [21], which recruited volunteers aged 40-69 years in 2006-10. Subsets of these participants (all ancestry) were invited to have brain (n∼50,000 analysed to date) and abdominal imaging (n∼15,000 analysed to date). UK Biobank received ethical approval from the Research Ethics Committee (reference 11/NW/0382), and all participants provided written informed consent. Participants with complete data were included. None had dementia.

### Alcohol consumption

Alcohol intake was self-reported at study baseline through a touchscreen questionnaire. Participants identified themselves as either current, never or previous drinkers. All groups were included if they had complete data on alcohol intake. For current drinkers, total weekly number of UK units (1 unit = ∼8g; a US standard drink is 14g) of alcohol consumed was calculated by summing across beverage types as previously described [22]. To compare directly associations with brain iron between alcohol intakes, weekly consumption was categorized into quintiles (and octiles for a sensitivity analysis). The lowest quintile of drinkers was used as the reference category to avoid under-estimating alcohol-related risks [23].

### Brain imaging

Participants underwent brain MRI at three imaging centres (Newcastle upon Tyne, Stockport or Reading) with identical Siemens Skyra 3T scanners (software VD13) using a standard 32-channel head coil (release date 02.02.2021). Susceptibility-weighted MRI-data (swMRI) were used for this study (3D GRE, TE1/TE2/TR=9.4/20/27ms, voxel size=0.8×0.8×2.0mm), as a measure sensitive to magnetic tissue constituents. Detailed image pre-processing and quality control pipelines are described elsewhere [24]. Two distinct and complementary metrics of brain iron deposition were used, T2* and quantitative susceptibility mapping (QSM), to produce image-derived phenotypes (IDPs). Whilst these metrics are coupled, consistent findings across the two will provide greater evidence that iron levels are affected. Subject-specific masks for fourteen sub-cortical regions were derived from the T1-weighted structural brain scan. We then calculated IDPs corresponding to the median T2* and χ values for each region. The fourteen regions correspond to left and right of seven subcortical structures. Two additional IDPs were calculated from QSM (left and right substantia nigra). In brief, T2* values were calculated from magnitude data. T2*-induced signal decay was calculated from the two echo times. T2* images were spatially filtered to reduce noise and transformed into the T1 space (by linear registration). QSM depends on phase images, which were obtained from individual coil channels, combined, masked and unwrapped. Magnetic susceptibility (χ) was calculated using a QSM pipeline including background field removal, dipole inversion and CSF referencing as described elsewhere [10]. Median χ values (in parts-per-billion) across voxels within each region were calculated, resulting in 16 QSM IDPs.

### Liver imaging

During the same visit that brain imaging was performed, some participants also underwent abdominal imaging on a Siemens 1.5T Magnetom Aera [25]. The acquisition used the LiverMultiScan protocol from Perspectum Diagnostics. A multi-echo spoiled gradient-echo single breath hold MRI sequence was acquired as a single transverse slice through the centre of the liver. Three ROI, representative of the liver parenchyma, were selected and mean T2* calculated. Iron levels were converted to mg/g. One IDP is liver fat, indexed using proton density fat fraction (PDFF, %), validated against MR spectroscopy and liver biopsy, and has a high specificity and sensitivity for non-alcoholic fatty liver disease [26, 27]. The other IDP, corrected T1 (cT1, milliseconds) was used as a marker of inflammation and fibrosis [28].

### Genetic variants

The distinct genetic architecture between different alcohol use traits motivates their separate analysis. Genetically predicted alcohol consumption was instrumented using 91/92 [depending on SNP or suitable proxy availability in outcome data] SNPs. These variants were associated with alcohol consumption (drinks per week at genome-wide significance; p<5×10^−8^) in the largest published GWAS comprising 941,280 individuals (GWAS & Sequencing Consortium of Alcohol and Nicotine) [29]. Alcohol use disorder (AUD) was instrumented using 24 conditionally independent GWS genetic variants in the largest published GWAS comprising the Million Veterans Program and the Psychiatric Genomics Consortium [30]. AUD cases were defined using ICD 9/10 codes within the MVP (n=45,995) and DSM-IV within the PGC (n=11,569). Genetic associations with serum markers of iron homeostasis (serum iron, ferritin, transferrin saturation (of iron-binding sites of transferrin occupied by iron), and total iron binding capacity) were obtained from the largest published GWAS to date (meta-analysis of deCODE, INTERVAL and Danish Blood Donor Study) [31]. Genetic associations with brain T2* and susceptibility (IV-outcomes) were obtained from the largest GWAS of brain imaging phenotypes [10, 32]. All genetic associations were based on GWAS of European ancestry samples.

### Clinical measures

We utilized the expanded cognitive battery performed on the subset of participants who underwent imaging, which included: trail-making tests (TMT A & B), prospective memory, and fluid intelligence. A larger subset of the wider UKB study (∼100k) underwent a subsequent TMT at online follow up. Motor function was assessed using simple reaction speed in the cognitive battery, handgrip strength and self-reported gait speed (slow/steady/brisk) at baseline.

### Statistical analysis

All analyses were performed in R (version 3.6.0).

### Observational

Separate linear regression models were used to assess the relationship between 1) alcohol consumption and, χ /T2*, 2) alcohol consumption and liver iron, 3) susceptibility and neuro-cognitive measures (ordered logistic regression for gait speed). Variables were quantile normalized to enforce Gaussianity in the data. Covariates previously associated with alcohol intake or iron levels and image-related confounders were included in the models: age, sex, imaging site, smoking status (never/previous/current), diabetes, body mass index, cholesterol, blood pressure and the full set of imaging confounds as recently proposed [33] which include imaging site, head motion, head position. For the subset with available data, dietary factors (meat, fish, bread, fruit and vegetable consumption) and dietary iron supplementation were also included as covariates, to test whether differences in diets according to alcohol intake were driving associations. Analyses were also controlled for genetic polymorphisms prevalent in European populations which affect iron absorption and metabolism and have been linked to brain phenotypes [34]. A risk score was calculated by multiplying the dosage of three SNPs by their associations with serum iron (betas from GWAS [35]): rs1800562 - in the HFE gene which is the main cause of hereditary hemochromatosis and carried by ∼5-10% of Europeans [36]; rs1799945 – also in HFE, carried by ∼15% Europeans; and rs855791 – which modulates hepicidin and ∼53% Europeans are heterozygotes. No included subjects reported iron chelator prescription. In a sensitivity analysis of females, menopause status at imaging was added as a covariate, as menstruation is protective against phenotypic expression of haemochromatosis [37].

For models with cognitive measures as the dependent variable, which are age-dependent, an age × iron interaction was included to explore whether iron altered age-related decline. Additional relevant covariates to cognition were included in these models: education, income, historical job type (Standard Occupational Classification 2000 [38]) and Townsend Deprivation Index. For models with χ /T2* as the dependent variable, an interaction between alcohol and liver iron was tested, to assess for synergistic neurotoxic effects of alcohol and iron. To adjust for multiple testing, Bonferroni and false discovery rate (FDR, 5%) corrected p values were calculated. Correction methods were separately applied to models testing susceptibility-alcohol associations and those testing susceptibility-cognitive associations.

Causal mediation analysis (CMA) was performed to understand the mechanisms by which alcohol impacts brain iron [39, 40]. Liver iron, reflecting systemic iron levels, was used as a mediator. The liver is the body’s primary iron store, and liver iron has better specificity for systemic iron stores than serum ferritin, which is an acute phase reactant liable to fluctuate according to inflammatory processes [15]. CMA tests the statistical significance of the direct from indirect (mediated) effects of alcohol on brain iron.

Two sets of *post-hoc* analyses were performed. First, given the prominence of basal ganglia iron levels in relation to alcohol we observed, associations with available motor phenotypes (simple reaction speed, handgrip strength and self-reported walking pace) were sought. Second, given observed associations between brain iron measures and age-related executive function, we assessed associations between alcohol consumption and executive function (at time of scan and online follow up). We could not examine relationships with relevant diseases, for example Parkinson’s due to insufficient numbers within the imaging sub study.

### Mendelian randomization

MR aims to estimate causal relationships using observational data [41]. Two sample linear MR was used to obtain estimates for the association between genetically predicted alcohol consumption and 1) brain susceptibility, 2) serum markers of iron homeostasis (as part of our investigation into the pathway by which alcohol influences brain iron). Analyses were conducted using *MendelianRandomization* (version 0.5.1) and *TwoSampleMR* (version 0.5.6) R packages. Variant harmonization ensured the association between SNPs and exposure and that between SNPs and the outcome reflected the same allele. Palindromic variants, where harmonization could not be confirmed, were excluded. Strands were aligned between studies. No proxies were used given the availability of SNPs across datasets. Several robust MR methods were performed to evaluate the consistency of the causal inference. Inverse variance weighted (IVW) analysis regresses the effect sizes of the variant-iron marker associations against the effect sizes of the variant-alcohol associations. The MR-Egger method uses a weighted regression with an unconstrained intercept to relax the assumption that all genetic variants are valid IVs (under the Instrument Strength Independent of Direct Effect (InSIDE) assumption) [42]. A non-zero intercept term can be interpreted as evidence of directional pleiotropy, where an instrument is independently associated with the outcome violating an MR assumption. The median and modal MR methods (reported in supplementary figures) are also more resistant to pleiotropy, as they are robust when up to 50% of genetic variants or more than not, respectively, are invalid. These methods are recommended in practice for sensitivity analyses as they require different assumptions to be satisfied, and therefore if estimates from such methods are similar, then any causal claim inferred is more credible. Analyses performed on one of the three cohorts (INTERVAL) meta-analysed in the serum iron GWAS was adjusted for alcohol consumption, so to check whether anti-conservative bias was impacting associations, a sensitivity analysis was run using weights derived solely from deCODE summary statistics which were unadjusted for alcohol.

## Results

Baseline characteristics of the included sample with complete data are shown in **Table 1**. Mean alcohol consumption was higher than current UK low risk guidelines (<14 units weekly), although within guidelines at the time for men (>21 units weekly pre-2016).

**Table 1:** Baseline characteristics of UK Biobank samples.

|  | Sample with brain imaging<br>N=22,254 | Sample with liver and brain<br>imaging<br>N= 11,024 |
| --- | --- | --- |
| Age<sup>1</sup>, years | 54.9 ± 7.4 | 54.9 ± 7.4 |
| Sex, n females (%) | 10821 (48.6) | 5788 (52.5) |
| Alcohol<sup>1</sup>, units weekly | 17.7 ± 16.0 | 17.0 ± 15.4 |
| Current smoker, n(%) | 1390 (6.2) | 665 (6.0) |
| Education, higher degree,<br>n(%) | 10819 (48.6) | 5029 (45.9) |
| Body mass index<sup>1</sup>, kg/m<sup>2</sup> | 26.5 ± 4.1 | 26.1 ± 3.9 |
| Systolic blood pressure<sup>1</sup>,<br>mmHg | 137.0 ± 18.6 | 136.9 ± 18.6 |

<sup>1</sup>Mean ± standard deviation

### Associations with brain iron

#### Observational analyses

0.09 standard deviation (S.D.) [95% confidence interval 0.07 to 0.10]), caudate (β=0.06 [0.04 to 0.07]) and substantia nigra (β=0.04 [0.02 to 0.05]), and lower iron in the thalami (β= -0.05 [-0.06 to -0.03 Alcohol consumption was associated with higher susceptibility, in bilateral putamen (beta = 0.09, 95% CI: 0.07 to 0.10), caudate (beta= 0.06, 95% CI: 0.04 to 0.07) and substantia nigra (beta=0.04, 95% CI: 0.02 to 0.05) **(Figure 2)**. Alcohol was associated with lower iron in the thalamus (beta= -0.05, 95% CI: -0.06 to - 0.03). Putamen iron levels were higher in those drinking 7 units (56g) weekly and greater compared to those drinking <7 units **(Figure 2(A)**). A sensitivity analysis with finer categorisation of alcohol intake confirmed that associations were not observed at lower intakes **(SFigure 1)**. Levels of drinking necessary to observe higher susceptibility values differed according to region **(SFigures 2-6)**. Observations between alcohol and susceptibility IDPs were evident at lower intakes in females **(SFigure 7)**. Menopause status did not associate with putamen susceptibility in females, nor did it not alter associations between alcohol and susceptibility **(STable 1)**. Inverse associations between thalamus susceptibility and alcohol were only observed in the heaviest drinking groups **(SFigures 5&6)**. In the subset with complete data, neither adjustment for dietary factors, including frequency of red meat consumption **(SFigure 8)**, nor dietary iron supplementation **(SFigure 9)** changed the pattern of associations. Findings with T2* were broadly consistent, although in more limited brain regions (putamen and caudate solely where correlation between T2* and susceptibility higher r= -0.58-0.75) **(SFigure 10)**.

**Figure 2:**
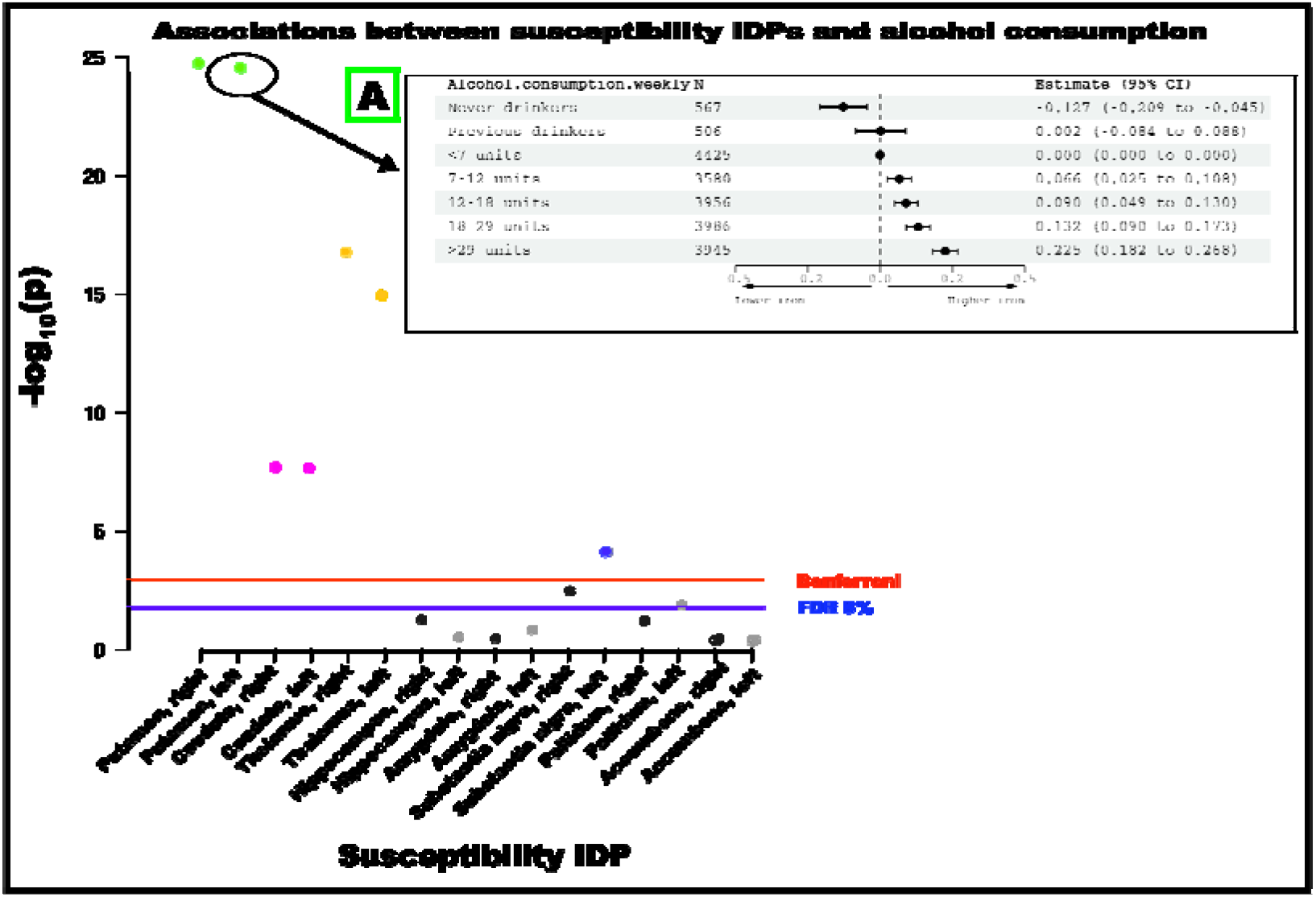
Associations between alcohol consumption and quantitative susceptibility mapping phenotypes for all participants. Insert **(A)** shows associations between alcohol intake (sample quintiles) and left putamen susceptibility where the reference group are those drinking <7 units (56g) weekly (reference, estimates (95% CI) set to 0). Estimates represent standard deviations and were generated from regression models adjusted for: age, sex, smoking, body mass index, diabetes, cholesterol, blood pressure, rs1800562, rs1799945 and rs855791 and full set of image-related confounds. Abbreviations: IDPs – imaging-derived phenotype, FDR – false discovery rate, CI – confidence interval.

#### Genetic analyses

Using 92 SNPs significantly associated with alcohol consumption, we found a positive association with left putamen susceptibility (IVW β=0.25, 95% CI: 0.01 to 0.49, p=0.04) and right hippocampus susceptibility (IVW β=0.28, 95% CI: 0.05 to 0.50, p=0.02)**(Figure 3)**. However, neither passed FDR correction for multiple comparisons. Although alternative methods gave consistent estimates, 95% confidence intervals were wide **(SFigure 11)**. Genetically-predicted AUD, instrumented with 24 SNPs, associated with higher right putamen susceptibility, although this did not survive multiple comparison correction (IVW β=0.18, 95% CI: 0.001 to 0.35, p=0.04). There were no other significant associations between genetically predicted AUD and other susceptibility measures **(STable 2)**.

**Figure 3:**
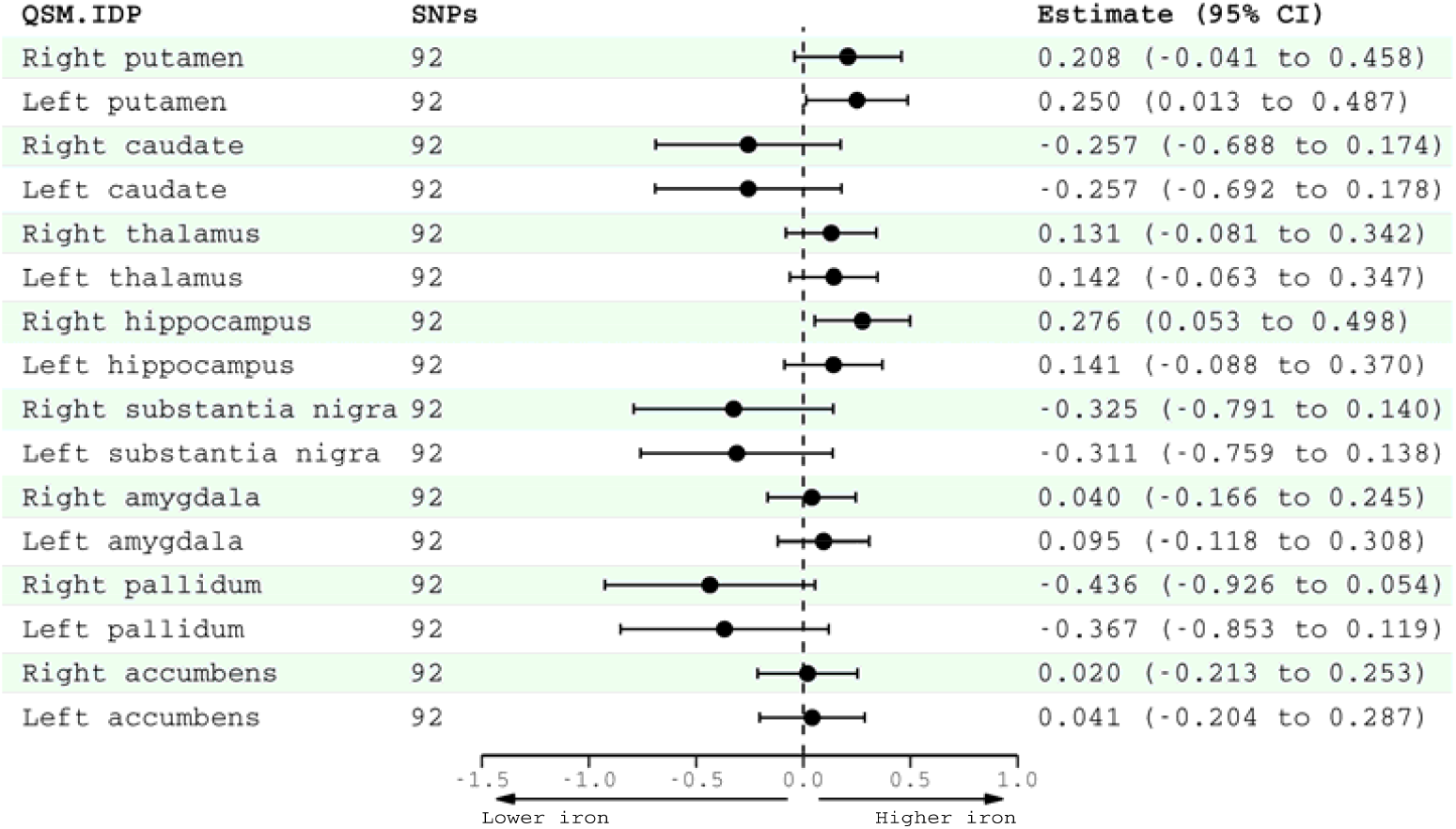
Two-sample linear Mendelian randomization estimates (inverse-variance weighted) for the association of genetically predicted alcohol consumption (GWAS & Sequencing Consortium of Alcohol and Nicotine) with quantitative susceptibility mapping image-derived phenotypes (UK Biobank). Effect estimates for alcohol consumption are per standard deviation increase in genetically-predicted log-transformed alcoholic drinks per week. Abbreviations: QSM IDP – quantitative susceptibility mapping imaging-derived phenotype, SNPs – single nucleotide polymorphism, CI – confidence interval.

### Pathways from alcohol to brain iron

Higher systemic iron levels (liver iron) were found to partially mediate alcohol’s association with putamen susceptibility in causal mediation analysis. 1 SD increase in weekly alcohol consumption was associated with a 0.05 (95% CI: 0.04-0.06) mg/g increase in liver iron (**Figure 4**). 1 mg/g increase in liver iron was associated with a 0.43 (95% CI: 0.35-0.51) SD increase in susceptibility. In this sample, 32[22-49] % of alcohol’s total effect on brain iron was mediated via higher systemic iron levels (the indirect effect). The other 68% is mediated via other pathways (the direct effect).

**Figure 4:**
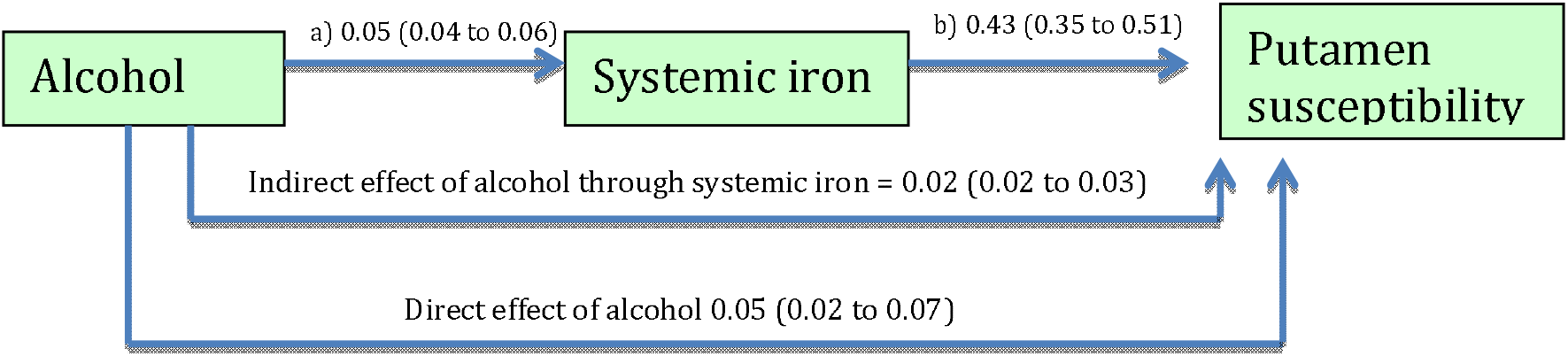
Mediation model of the association of alcohol and increased putamen iron (measured by susceptibility, quantile normalized). N=8936 UK Biobank participants. Numbers on the arrows are regression coefficients (with 95% CIs) reflecting: a) Change in liver iron (mg/g) for a 1 unit increase in alcohol intake weekly, b) Change in susceptibility (standard deviation (95% confidence intervals)) for a 1mg/g increase in liver iron. The indirect and direct effects (standardized) derived from the mediation analysis are reported on the arrows linking alcohol to susceptibility. Models were adjusted for: age, sex, imaging site, blood pressure, body mass index, cholesterol, smoking, and polygenic risk score for serum iron.

Alcohol consumption (>11 units) was associated with higher liver iron measured by MRI, a robust marker of systemic iron stores (**SFigure 12**). This is within currently defined UK ‘low risk’ guidelines (<14 units weekly). Iron appeared to be a more sensitive liver marker of alcohol consumption than fat (PDFF) or fibrosis (cT1) (**SFigures 13&14**).

Using 24 SNPs significantly associated with AUD, there were statistically significant associations between genetically predicted AUD and serum iron (IVW β =0.12, 95% CI: 0.05 to 0.19, p=0.001) as well as transferrin saturation (IVW β =0.11, 95% CI: 0.03 to 0.12, p=0.006) that survive even stringent Bonferroni multiple testing correction (**Figure 5**). Confidence intervals for MR Egger estimates were larger (serum iron β=0.17, 95% CI: 0.009 to 0.34; transferrin saturation β=0.16, 95% CI: -0.02 to 0.33) but broadly consistent (**SFigure 15**). Although estimates for associations between genetically–predicted alcohol consumption (instrumented by 91 SNPs) and serum iron/transferrin saturation were of a similar magnitude, confidence intervals were wider. In sensitivity analyses solely using data from deCODE which did not adjust their GWAS for alcohol, overall findings were unchanged (**SFigure 16**).

**Figure 5:**
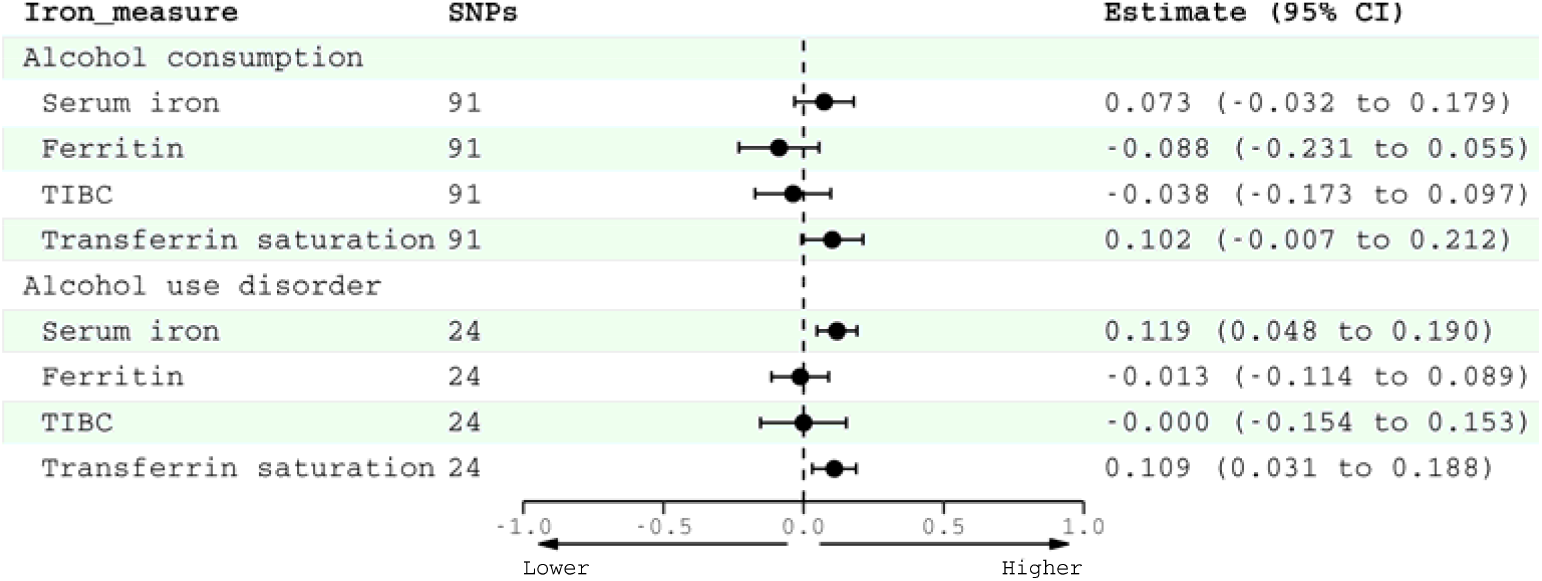
Mendelian randomization estimates (two-sample design) for the association of genetically predicted alcohol consumption (GWAS & Sequencing Consortium of Alcohol and Nicotine) and alcohol use disorder (Million Veterans Program & Psychiatric Genomics Consortium) with serum iron markers (deCODE, INTERVAL and Danish Blood Donor Study). Inverse-variance weighted estimates are shown. Effect estimates for alcohol consumption are per SD increase in genetically-predicted log-transformed alcoholic drinkers per week, and for AUD having a diagnosis of AUD. Abbreviations: TIBC – total iron-binding capacity, SNPs – single nucleotide polymorphisms, CI – confidence intervals.

### Clinical relevance of elevated brain iron

Higher putamen, caudate and hippocampal susceptibility were associated with greater age-related slowing of executive function, and higher putamen and caudate susceptibility with greater age-related differences in fluid intelligence (**Figure 6 (A-C) & STable 3**). Interactions between age and putamen susceptibility were significant in the models fitting TMT A duration (β=0.005, 95% CI: 0.003 to 0.007), TMT B duration (β=0.004, 95% CI: 0.002 to 0.006) and fluid intelligence (β=-0.004, 95% CI: -0.006 to -0.002). Caudate susceptibility significantly interacted with age in models with TMT A (β=2.87×10^−3^, 95% CI: 7.66×10^−4^ to 4.97×10^−3^), TMT B (β=3.37e-3, 95% CI: 1.25×10^−3^ to 5.49×10^−3^) and fluid intelligence (β= -2.33×10^−3^, 95% CI: -4.47×10^−3^ to -1.93×10^−4^). Higher right hippocampus susceptibility associated with greater age-related increases in TMT A duration (β= 3.96×10^−3^, 95% CI: 8.43×10^−4^ to 5.05×10^−3^). Neither substantia nigra nor thalamus susceptibility was associated with age-related differences in measured cognitive function.

**Figure 6:**
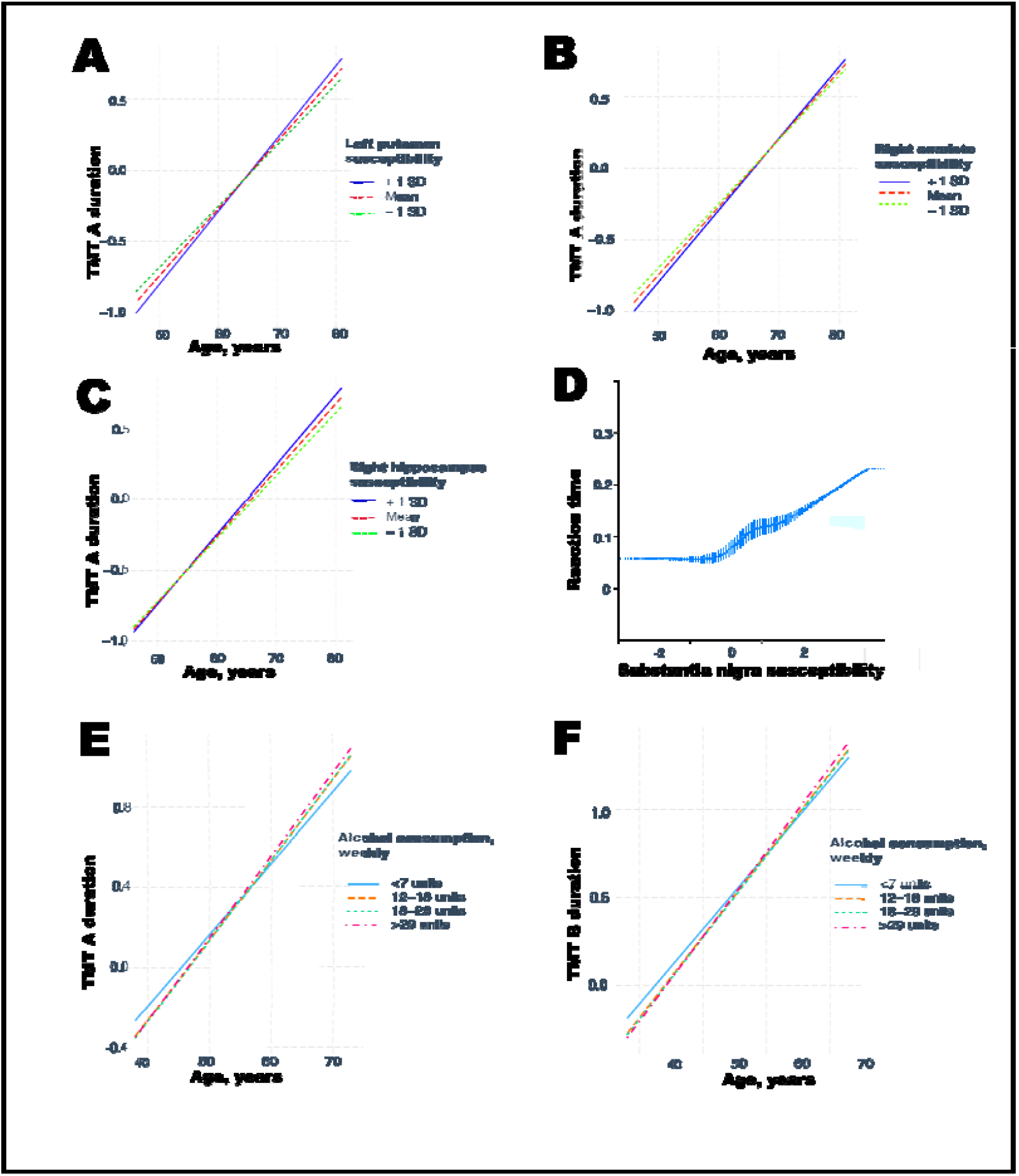
**(A)** Age-related executive function (at time of scan) according to putamen susceptibility. Trail-making test (TMT) durations and susceptibility are quantile normalized. **(B)** Age-related executive function according to caudate susceptibility. (C) Age-related executive function according to hippocampus susceptibility. **(D)** Reaction time (quantile normalized and fitted with restricted cubic spline, 5 knots) according to substantia nigra susceptibility. **(E) & (F)** Age-related executive function (at online follow up) according to alcohol consumption (quintiles) in weekly units. Only intakes significantly different from reference group (<7 units) are plotted for clearer visualisation. Graphs generated from regression models controlled for: age, sex, imaging site, diabetes, smoking, income, education, body mass index, blood pressure, cholesterol, historical job type.

Higher right substantia nigra susceptibility was associated with slower simple reaction time (β= 0.002, 95% CI: 0.009 to 0.04) but not with other motor deficits (**STable 4**). However there were no significant associations between putamen, caudate, hippocampus or thalamus susceptibility and simple reaction time at the time of scan, or with age-related reaction time differences (**STable 3**). There were no significant associations between susceptibility measures and self-reported walking pace (OR=1.00, 95% CI: 0.97-1.03).

Grip strength was positively associated with putamen (β= 1.74×10^−2^, 95% CI: 5.19×10^−3^ to 2.96×10^−2^) and caudate (β= 1.55×10^−2^, 95% CI: 3.71×10^−3^ to 2.74×10^−2^) susceptibility, although caudate associations would not survive multiple testing comparison (**STable 4**).

In a *post-hoc* analysis, acceleration of age-related decline in executive function was evident in those drinking >12 units weekly (12-18 vs. <7 units weekly β= - 0.24, 95% CI: -0.40 to -0.09) (**Figure 6 (E) & (F)**). However, this was only observed within the larger sample that participated in the later online follow up, preventing exploration of whether brain iron was mediating this pathway.

## Discussion

### Key findings

Alcohol consumption, including at low levels, was associated with higher brain iron in multiple basal ganglia regions. There was some evidence to support a causal effect of alcohol on the putamen and hippocampus, although this did not survive multiple testing correction. Alcohol was associated with both higher liver iron, an index of systemic iron load, and AUD causally related to higher serum iron markers. Brain iron accumulation in drinkers was only partially mediated via higher systemic iron. Markers of higher brain iron (higher susceptibility) were associated with greater age-related executive function and fluid intelligence differences, and slower reaction speed.

### Results in context of other literature

To our knowledge, this is the largest study of moderate alcohol consumption and multi-organ iron accumulation. It is also the first study to use MR to investigate causality of alcohol on serum and brain iron.

The accumulation of iron in the brain we observed in moderate drinkers overlaps with findings of an observational study in AUD. Higher putamen and caudate iron levels were described in a small study of males with AUD (n=20) [9]. These individuals were drinking substantially more than our sample – a mean of 22 standard drinks per day (>37 units daily). In contrast, we observed associations in those drinking just >7 units per week. A recent phenomewide association study of quantitative susceptibility in the same dataset reported significant associations in basal ganglia regions with higher frequency binge-drinking [10]. Regional heterogeneity in iron concentrations is well described although the aetiology is not understood [43]. The basal ganglia, including the putamen [44], have some of the highest iron concentrations in the brain and suffer the greatest age-related increases [45]. In this cohort, associations with susceptibility and T2* measures were observed at lower alcohol intakes in females. In haemochromatosis females are relatively protected against the clinical manifestations of iron overload through menstrual blood loss [46]. The majority of our included sample however (70%) was post-menopausal and menopause status did not alter alcohol-brain iron associations. Sex differences in alcohol metabolism therefore may be responsible. These findings do not support current UK ‘low risk’ drinking guidelines which recommend identical amounts for males and females [47]. We found some support for a causal relationship between alcohol consumption and susceptibility in the putamen and hippocampus, and between AUD and putamen susceptibility in MR analysis. Although these associations did not survive multiple comparisons correction they are in the same direction as the highly significant observational associations. Associations between genetically predicted alcohol and susceptibility in other regions were not significant. We suspect this results from our limited power to detect small associations despite the sample size, given that the genetic instruments explain less than 1% of the phenotypic variation in alcohol consumption [29].

Our MR results provide evidence for a causal role of AUD in increasing serum iron and transferrin saturation, a sensitive marker of iron overload [48]. Whilst genetically predicted alcohol use was not significantly associated with ferritin, this mirrors findings in early haemochromatosis, where ferritin levels can be normal and transferrin saturation is the earliest marker of iron overload [49]. The associations we found with liver iron, a reliable marker of systemic iron stores, were consistent with the serum results. In fact, in our study, which we believe the largest investigation of alcohol and liver iron by an order of magnitude [50], iron levels were the most sensitive liver marker of alcohol-related damage. Alcohol suppresses hepcidin production, the major hormone regulating iron homeostasis [51]. This suppression increases intestinal absorption of dietary iron [52] and limits export of iron from hepatocytes. In our causal mediation analysis, higher systemic iron levels only explained 32% of alcohol’s effects on brain iron, suggesting other mechanisms are also involved. These could include an increase of blood-brain barrier permeability to iron, in turn mediated by reduced thiamine which commonly occurs in AUD due to a combination of inadequate dietary intake, reduced absorption and metabolic changes [53, 54]. In cerebral automosal dominant arteriopathy with subcortical infarcts and leukoencephalopathy (CADASIL) patients, iron leakage has been linked to blood-brain barrier permeability [55]. Other possible mechanisms include dopamine surges following alcohol ingestion or chronic inflammatory processes [56].

Higher putamen and caudate susceptibility were associated with steeper age-related drops in executive function and fluid intelligence, but not with simple motor tasks. Most, but not all, previous work has highlighted the importance of the putamen to complex motor tasks [57]. Interestingly, both trail making and the fluid intelligence tasks were performed within a time limit, and perhaps represent a measure of motor response linked to cognition, rather than a simple motor response. Trail-making tests appear to be amongst the most sensitive to aging effects in the UKB cognitive battery [58]. Frontal dysfunction is well described in chronic heavy alcohol use [59]. Several putamen metrics have been associated with executive function, including blood flow [60], structural atrophy [61] and functional connectivity [62]. Iron accumulation in the putamen has also been described in developmental stuttering [63] and CADASIL [64]. How iron deposition could result in cognitive deficits requires further investigation. Iron co-localises in the brain with tau and beta amyloid [65], and can cause apoptosis and ferroptosis [66]. Higher substantia nigra susceptibility associated with slower reaction speed. The substantia nigra plays a vital role in movement regulation, and iron deposition in the substantia nigra has been linked to Parkinson’s disease [67, 68], a disorder with marked impairments in reaction speed [69].

We did not observe widespread associations between susceptibility or T2* and other cognitive tests or self-reported motor measures. Brain iron is likely to be an early marker of disease, and participants may have been examined too early in the process to detect clinical manifestations. Additionally, we are not likely to have captured the best phenotypes to assess basal ganglia function in the absence of objective motor measurements such as gait speed or a pegboard test. Self-reported walking speed may poorly approximate actual motor function. The cognitive tests were limited in scope and concerns have been raised about the reliability of the tests used [58]. Healthy selection biases in UKB are well described, and are likely magnified in the imaging subsample, but will equally bias the study towards null-results [70]. Furthermore, associations in UKB seem to track with those observed in representative cohorts [71].

### Clinical implications

Never drinkers appeared to have the lowest levels of brain iron. This is in keeping with our earlier work indicating there may be no safe level of alcohol consumption for brain health [22]. There are major public health implications given the prevalence of moderate drinking. If elevated brain iron is confirmed as a mechanism by which alcohol leads to cognitive decline, there are opportunities for intervention. Iron chelation therapy is already being investigated for Alzheimer’s and Parkinson’s diseases [17, 18, 72]. Furthermore, if reduced thiamine is mediating brain iron accumulation, then interventions to improve nutrition and thiamine supplementation could be extended beyond harmful and dependent drinkers, as is currently recommended [73], to moderate drinkers.

### Limitations

Changes in T2* and χ can reflect changes in iron but also myelin [74, 75]. One key difference between T2* and χ is that iron (paramagnetic) and myelin (diamagnetic) have the opposite effect on χ in QSM, but the same effect on T2*. Hence the positive associations we observed between χ and alcohol could theoretically be driven by increased iron or reduced myelin. If the latter, then alcohol would also be positively associated with T2* (reduced myelin leads to longer T2*). In contrast, we observed negative associations between T2* and alcohol. This supports our interpretation that increased iron is driving our results, given one highly plausible assumption, that alcohol does not increase grey matter myelination [76, 77].

Partial volume effects could confound associations between hippocampal susceptibility and alcohol. For example, hippocampal atrophy, previously observed in drinkers [1], could be conflated with changes in χ. However this would tend to reduce estimated χ. Alcohol was self-reported, but this is the only feasible method to ascertain intake at scale. Serum markers of iron homeostasis were not directly measured in UKB. Although analyses were controlled for the strongest SNPs associated with serum iron, these are likely to explain a low proportion of the variance. MR techniques rely on a number of assumptions which we have tried to test where possible, but residual uncertainty inevitably remains. Liver T2* has been useful in some studies to monitor iron overload, but further validation of this biomarker as a diagnostic marker of iron overload is needed [78]. Genetic variants explain a low variance of alcohol traits. Therefore our analysis within the imaging sample, despite its large size, was not powered to detect small associations with alcohol. Mediation analysis is not experimental in design, and relies on intervention-outcome, intervention-mediator, and mediator-outcome effects being unconfounded to permit valid causal inferences. In this study liver and brain iron were measured at the same time, meaning reverse causation is possible. However, it is difficult to conceive of a plausible mechanism by which brain iron levels could substantially affect systemic iron.

## Conclusions

In this large sample of UKB participants, we find evidence for elevated susceptibility and reduced T2* in basal ganglia regions with even moderate alcohol consumption. These changes likely reflect increased iron concentrations. Alcohol-related brain iron may be partially mediated by higher systemic iron levels, but it is likely there are additional mechanisms involved. Greater age-related decreases in executive function and fluid intelligence, and slower reaction speeds were seen with markers of higher basal ganglia iron. Brain iron accumulation is a plausible mechanism for alcohol-related cognitive decline.

## Data Availability

Data is available through a successful UK Biobank application.

## Acknowledgements

This work was undertaken using UK Biobank application numbers 55929 & 8107. AT is supported by a Wellcome Trust (https://wellcome.org/) fellowship (216462/Z/19/Z).

CW is funded, in part, by the China Scholarship Council (CSC, https://www.chinesescholarshipcouncil.com/). KPE is funded by the UK Medical Research Council (https://mrc.ukri.org/, G1001354 & MR/K013351/1) and the European Commission (https://ec.europa.eu/programmes/horizon2020/en/home, Horizon 2020 732592). CM is supported by the Oxford NIHR Biomedical Research Centre and the Oxford BHF Centre of Research Excellence. JG, DL and HZ are supported by the US Department of Veterans Affairs (https://www.research.va.gov/funding/, I01CX001849). SB is supported by a Sir Henry Dale Fellowship jointly funded by the Wellcome Trust and the Royal Society (https://royalsociety.org/, 204623/Z/16/Z). S.Bell was supported by the British Heart Foundation (https://www.bhf.org.uk/for-professionals/information-for-researchers/what-we-fund, RG/16/4/32218) and the NIHR Cambridge Biomedical Research Centre (BRC-1215-20014). MH is supported by the Wellcome Trust and NIHR Oxford Biomedical Research Centre. SS is supported by a Wellcome Trust Collaborative Award 215573/Z/19/Z. KLM is supported by a Wellcome Trust Senior Research Fellowship 202788/Z/16/Z. This work was also supported by the Li Ka Shing Centre for Health Information and Discovery, NIH grant (https://www.nih.gov/, TN: R01EB026859), the National Institute for Health Research Cambridge Biomedical Research Centre (BRC-1215-20014), and a Wellcome Trust award (TN: 100309/Z/12/Z).

The views expressed are those of the author(s) and not necessarily those of the NIHR or the Department of Health and Social Care. The funding organisations had no role in any of the following: design and conduct of the study; collection, management, analysis, and interpretation of the data; review, or approval of the manuscript.

## References

1. Topiwala A, Allan CL, Valkanova V, Zsoldos E, Filippini N, Sexton C, et al. Moderate alcohol consumption as risk factor for adverse brain outcomes and cognitive decline: longitudinal cohort study. bmj. 2017;357.

2. Mukamal KJ, Kuller LH, Fitzpatrick AL, Longstreth Jr WT, Mittleman MA, Siscovick DS. Prospective study of alcohol consumption and risk of dementia in older adults. Jama. 2003;289(11):1405–13.

3. Organization WH. Global status report on alcohol and health 2018: executive summary. World Health Organization; 2018.

4. Visontay R, Rao RT, Mewton L. Alcohol use and dementia: new research directions. Current Opinion in Psychiatry. 2021;34(2):165–70.

5. Listabarth S, König D, Vyssoki B, Hametner S. Does thiamine protect the brain from iron overload and alcohol-related dementia? Alzheimer’s & Dementia. 2020;16(11):1591–5.

6. Nielsen J, Jensen LN, Krabbe K. Hereditary haemochromatosis: a case of iron accumulation in the basal ganglia associated with a parkinsonian syndrome. Journal of Neurology, Neurosurgery & Psychiatry. 1995;59(3):318–21.

7. Lane DJ, Ayton S, Bush AI. Iron and Alzheimer’s disease: an update on emerging mechanisms. Journal of Alzheimer’s Disease. 2018;64(1):S379–S95.

8. Oakley A, Collingwood J, Dobson J, Love G, Perrott H, Edwardson J, et al. Individual dopaminergic neurons show raised iron levels in Parkinson disease. Neurology. 2007;68(21):1820–5.

9. Juhás M, Sun H, Brown MR, Mackay MB, Mann KF, Sommer WH, et al. Deep grey matter iron accumulation in alcohol use disorder. Neuroimage. 2017;148:115–22.

10. Wang C, Martins-Bach AB, Alfaro-Almagro F, Douaud G, Klein JC, Llera A, et al. Phenotypic and genetic associations of quantitative magnetic susceptibility in UK Biobank brain imaging. bioRxiv. 2021.

11. Merryweather-Clarke AT, Pointon JJ, Shearman JD, Robson K. Global prevalence of putative haemochromatosis mutations. Journal of medical genetics. 1997;34(4):275–8.

12. Whitfield JB, Zhu G, Heath AC, Powell L, Martin N. Effects of alcohol consumption on indices of iron stores and of iron stores on alcohol intake markers. Alcoholism: Clinical and Experimental Research. 2001;25(7):1037–45. 13.

13. Duane P, Raja K, Simpson R, Peters T. Intestinal iron absorption in chronic alcoholics. Alcohol and Alcoholism. 1992;27(5):539–44.

14. Ioannou GN, Dominitz JA, Weiss NS, Heagerty PJ, Kowdley KV. The effect of alcohol consumption on the prevalence of iron overload, iron deficiency, and iron deficiency anemia. Gastroenterology. 2004;126(5):1293–301.

15. Barry M, Sherlock S. Measurement of liver-iron concentration in needle-biopsy specimens. The Lancet. 1971;297(7690):100–3.

16. Brittenham GM, Farrell DE, Harris JW, Feldman ES, Danish EH, Muir WA, et al. Magnetic-susceptibility measurement of human iron stores. New England Journal of Medicine. 1982;307(27):1671–5.

17. Bagwe-Parab S, Kaur G. Molecular targets and therapeutic interventions for iron induced neurodegeneration. Brain research bulletin. 2020;156:1–9.

18. Martin-Bastida A, Ward RJ, Newbould R, Piccini P, Sharp D, Kabba C, et al. Brain iron chelation by deferiprone in a phase 2 randomised double-blinded placebo controlled clinical trial in Parkinson’s disease. Scientific reports. 2017;7(1):1–9.

19. De Barros A, Arribarat G, Combis J, Chaynes P, Péran P. Matching ex vivo MRI with iron histology: pearls and pitfalls. Frontiers in neuroanatomy. 2019;13:68.

20. Wood JC, Zhang P, Rienhoff H, Abi-Saab W, Neufeld EJ. Liver MRI is more precise than liver biopsy for assessing total body iron balance: a comparison of MRI relaxometry with simulated liver biopsy results. Magnetic resonance imaging. 2015;33(6):761–7.

21. Bycroft C, Freeman C, Petkova D, Band G, Elliott LT, Sharp K, et al. The UK Biobank resource with deep phenotyping and genomic data. Nature. 2018;562(7726):203–9.

22. Topiwala A, Ebmeier KP, Maullin-Sapey T, Nichols TE. No safe level of alcohol consumption for brain health: observational cohort study of 25,378 UK Biobank participants. medRxiv. 2021.

23. Naimi T, Chikritzhs T, Stockwell T. Commentary on Di Castelnuovo et al: Implications of using low volume drinkers instead of never drinkers as the reference group. Addiction. 2022;117(2):327–9.

24. Alfaro-Almagro F, Jenkinson M, Bangerter NK, Andersson JL, Griffanti L, Douaud G, et al. Image processing and Quality Control for the first 10,000 brain imaging datasets from UK Biobank. Neuroimage. 2018;166:400–24.

25. Mckay A, Wilman HR, Dennis A, Kelly M, Gyngell ML, Neubauer S, et al. Measurement of liver iron by magnetic resonance imaging in the UK Biobank population. PloS one. 2018;13(12):e0209340.

26. Gu J, Liu S, Du S, Zhang Q, Xiao J, Dong Q, et al. Diagnostic value of MRI-PDFF for hepatic steatosis in patients with non-alcoholic fatty liver disease: a meta-analysis. European radiology. 2019;29(7):3564–73.

27. Reeder SB, Pineda AR, Wen Z, Shimakawa A, Yu H, Brittain JH, et al. Iterative decomposition of water and fat with echo asymmetry and least-squares estimation (IDEAL): application with fast spin-echo imaging. Magnetic Resonance in Medicine: An Official Journal of the International Society for Magnetic Resonance in Medicine. 2005;54(3):636–44.

28. Pavlides M, Banerjee R, Sellwood J, Kelly CJ, Robson MD, Booth JC, et al. Multiparametric magnetic resonance imaging predicts clinical outcomes in patients with chronic liver disease. Journal of hepatology. 2016;64(2):308–15. 29.

29. Liu M, Jiang Y, Wedow R, Li Y, Brazel DM, Chen F, et al. Association studies of up to 1.2 million individuals yield new insights into the genetic etiology of tobacco and alcohol use. Nature genetics. 2019;51(2):237–44.

30. Zhou H, Sealock JM, Sanchez-Roige S, Clarke T-K, Levey DF, Cheng Z, et al. Genome-wide meta-analysis of problematic alcohol use in 435,563 individuals yields insights into biology and relationships with other traits. Nature neuroscience. 2020;23(7):809–18.

31. Bell S, Rigas AS, Magnusson MK, Ferkingstad E, Allara E, Bjornsdottir G, et al. A genome-wide meta-analysis yields 46 new loci associating with biomarkers of iron homeostasis. Communications biology. 2021;4(1):1–14.

32. Smith SM, Douaud G, Chen W, Hanayik T, Alfaro-Almagro F, Sharp K, et al. An expanded set of genome-wide association studies of brain imaging phenotypes in UK Biobank. Nature neuroscience. 2021;24(5):737–45.

33. Alfaro-Almagro F, Mccarthy P, Afyouni S, Andersson JL, Bastiani M, Miller KL, et al. Confound modelling in UK Biobank brain imaging. NeuroImage. 2021;224:117002.

34. Atkins JL, Pilling LC, Heales CJ, Savage S, Kuo C-L, Kuchel GA, et al. Hemochromatosis Mutations, Brain Iron Imaging, and Dementia in the UK Biobank Cohort. Journal of Alzheimer’s Disease. 2021(Preprint):1-9.

35. Benyamin B, Esko T, Ried JS, Radhakrishnan A, Vermeulen SH, Traglia M, et al. Novel loci affecting iron homeostasis and their effects in individuals at risk for hemochromatosis. Nature communications. 2014;5(1):1–11.

36. Medicine NLO. Reference SNP (rs) report. 2021.

37. Brissot P, Pietrangelo A, Adams PC, De Graaff B, Mclaren CE, Loréal O. Haemochromatosis. Nature Reviews Disease Primers. 2018;4(1):1–15.

38. Statistics OON. Standard Occupational Classification 2000: SOC 2000. 2000.

39. Imai K, Keele L, Tingley D, Yamamoto T. Causal mediation analysis using R. Advances in social science research using R: Springer; 2010. p. 129–54.

40. Lee H, Herbert RD, Mcauley JH. Mediation analysis. Jama. 2019;321(7):697–8.

41. Smith GD, Ebrahim S. Mendelian randomization: prospects, potentials, and limitations. International journal of epidemiology. 2004;33(1):30–42.

42. Bowden J, Davey Smith G, Burgess S. Mendelian randomization with invalid instruments: effect estimation and bias detection through Egger regression. International journal of epidemiology. 2015;44(2):512–25.

43. Ramos P, Santos A, Pinto NR, Mendes R, Magalhães T, Almeida A. Iron levels in the human brain: a post-mortem study of anatomical region differences and age-related changes. Journal of Trace Elements in Medicine and Biology. 2014;28(1):13–7.

44. Mcallum EJ, Hare DJ, Volitakis I, Mclean CA, Bush AI, Finkelstein DI, et al. Regional iron distribution and soluble ferroprotein profiles in the healthy human brain. Progress in neurobiology. 2020;186:101744.

45. Bilgic B, Pfefferbaum A, Rohlfing T, Sullivan EV, Adalsteinsson E. MRI estimates of brain iron concentration in normal aging using quantitative susceptibility mapping. Neuroimage. 2012;59(3):2625–35.

46. Pilling LC, Tamosauskaite J, Jones G, Wood AR, Jones L, Kuo C-L, et al. Common conditions associated with hereditary haemochromatosis genetic variants: cohort study in UK Biobank. bmj. 2019;364.

47. Care DOHaS. UK chief medical officers’ guidelines on how to keep health risks from drinking alcohol to a low level. 2016.

48. Elsayed M, Sharif M, Stack A. Transferrin saturation: a body iron biomarker. Advances in clinical chemistry. 2016;75:71–97.

49. Mclaren CE, Mclachlan GJ, Halliday JW, Webb SI, Leggett BA, Jazwinska EC, et al. Distribution of transferrin saturation in an Australian population: relevance to the early diagnosis of hemochromatosis. Gastroenterology. 1998;114(3):543–9.

50. Schutte R, Papageorgiou M, Najlah M, Huisman HW, Ricci C, Zhang J, et al. Drink types unmask the health risks associated with alcohol intake–prospective evidence from the general population. Clinical Nutrition. 2020;39(10):3168–74. 51.

51. Ohtake T, Saito H, Hosoki Y, Inoue M, Miyoshi S, Suzuki Y, et al. Hepcidin is down-regulated in alcohol loading. Alcoholism: Clinical and Experimental Research. 2007;31:S2–S8.

52. Ganz T, Nemeth E. Hepcidin and iron homeostasis. Biochimica et Biophysica Acta (BBA)-Molecular Cell Research. 2012;1823(9):1434–43.

53. Harata N, Iwasaki Y. Evidence for early blood-brain barrier breakdown in experimental thiamine deficiency in the mouse. Metabolic brain disease. 1995;10(2):159–74.

54. Martin PR, Singleton CK, Hiller-Sturmhöfel S. The role of thiamine deficiency in alcoholic brain disease. Alcohol research & health. 2003;27(2):134. 55.

55. Uchida Y, Kan H, Sakurai K, Arai N, Inui S, Kobayashi S, et al. Iron leakage owing to blood–brain barrier disruption in small vessel disease CADASIL. Neurology. 2020;95(9):e1188–e98.

56. Yoder KK, Constantinescu CC, Kareken DA, Normandin MD, Cheng TE, O’connor SJ, et al. Heterogeneous effects of alcohol on dopamine release in the striatum: a PET study. Alcoholism: Clinical and Experimental Research. 2007;31(6):965–73.

57. Delong M. Putamen: activity of single units during slow and rapid arm movements. Science. 1973;179(4079):1240–2.

58. Fawns-Ritchie C, Deary IJ. Reliability and validity of the UK Biobank cognitive tests. PloS one. 2020;15(4):e0231627.

59. Kopelman MD. Frontal dysfunction and memory deficits in the alcoholic Korsakoff syndrome and Alzheimer-type dementia. Brain. 1991;114(1):117–37. 60.

60. O’carroll R.E. HPC, Ebmeier K.P., Dougall N., Murray C, Best Jjk, Bouchier Iad, Goodwin Gm. Regional cerebral blood flow and cognitive function in patients with chronic liver disease. The Lancet. 1991;337(May 25).

61. Li P, Wang Y, Jiang Y, Chen X, Dong Q, Cui M. Association between cognition and gait in elderly people: The putamen as a shared substrate: Neuropsychology/Neuropsychological correlates of physiologic markers of cognitive decline/Dementia. Alzheimer’s & Dementia. 2020;16:e041905.

62. Fjell AM, Sneve MH, Grydeland H, Storsve AB, Walhovd KB. The disconnected brain and executive function decline in aging. Cerebral cortex. 2017;27(3):2303–17.

63. Cler GJ, Krishnan S, Papp D, Wiltshire CE, Chesters J, Watkins KE. Elevated iron concentration in putamen and cortical speech motor network in developmental stuttering. Brain. 2021;144(10):2979–84.

64. Sun C, Wu Y, Ling C, Xie Z, Kong Q, Fang X, et al. Deep gray matter Iron deposition and its relationship to clinical features in cerebral autosomal dominant Arteriopathy with subcortical infarcts and leukoencephalopathy patients: a 7.0-T magnetic resonance imaging study. Stroke. 2020;51(6):1750–7.

65. Van Bergen J, Li X, Hua J, Schreiner S, Steininger S, Quevenco F, et al. Colocalization of cerebral iron with amyloid beta in mild cognitive impairment. Scientific reports. 2016;6(1):1–9.

66. Dixon SJ, Lemberg KM, Lamprecht MR, Skouta R, Zaitsev EM, Gleason CE, et al. Ferroptosis: an iron-dependent form of nonapoptotic cell death. Cell. 2012;149(5):1060–72.

67. Gorell J, Ordidge R, Brown G, Deniau J, Buderer N, Helpern J. Increased iron-related MRI contrast in the substantia nigra in Parkinson’s disease. Neurology. 1995;45(6):1138–43.

68. Sofic E, Paulus W, Jellinger K, Riederer P, Youdim M. Selective increase of iron in substantia nigra zona compacta of parkinsonian brains. Journal of neurochemistry. 1991;56(3):978–82.

69. Cooper JA, Sagar HJ, Tidswell P, Jordan N. Slowed central processing in simple and go/no-go reaction time tasks in Parkinson’s disease. Brain. 1994;117(3):517–29.

70. Fry A, Littlejohns TJ, Sudlow C, Doherty N, Adamska L, Sprosen T, et al. Comparison of sociodemographic and health-related characteristics of UK Biobank participants with those of the general population. American journal of epidemiology. 2017;186(9):1026–34.

71. Batty GD, Gale CR, Kivimäki M, Deary IJ, Bell S. Comparison of risk factor associations in UK Biobank against representative, general population based studies with conventional response rates: prospective cohort study and individual participant meta-analysis. bmj. 2020;368.

72. Singh YP, Pandey A, Vishwakarma S, Modi G. A review on iron chelators as potential therapeutic agents for the treatment of Alzheimer’s and Parkinson’s diseases. Molecular diversity. 2019;23(2):509–26.

73. Nice. Alcohol-use disorders: diagnosis and management of physical complications. 2017.

74. Stüber C, Morawski M, Schäfer A, Labadie C, Wähnert M, Leuze C, et al. Myelin and iron concentration in the human brain: a quantitative study of MRI contrast. Neuroimage. 2014;93:95–106.

75. Duyn JH, Schenck J. Contributions to magnetic susceptibility of brain tissue. NMR in Biomedicine. 2017;30(4):e3546.

76. Kilpatrick LA, Alger JR, O’neill J, Joshi SH, Narr KL, Levitt JG, et al. Impact of prenatal alcohol exposure on intracortical myelination and deep white matter in children with attention deficit hyperactivity disorder. Neuroimage: Reports. 2022;2(1):100082.

77. Ozer E, Sarioglu S, Güre A. Effects of prenatal ethanol exposure on neuronal migration, neuronogenesis and brain myelination in the mice brain. Clinical neuropathology. 2000;19(1):21–5.

78. Sarigianni M LA, Vlachaki E, Paschos P, Athanasiadou E, Montori Vm, Murad Mh, Tsapas A. Accuracy of magnetic resonance imaging in diagnosis of liver iron overload: a systematic review and meta-analysis. Gastroenterol Hepatol. 2015;13(1):55–63.

