## Supplementary tables for "Impact of moderate alcohol consumption on brain iron and cognition: observational and genetic analyses"

|  | **Association with left putamen susceptibility** |
| --- | --- |
| **Post- vs. pre-menopausal** | -7.96e-2 (-1.66e-1 to 0.01) |
| **Hysterectomy vs. pre-menopausal** | -9.68e-2 (-2.01e-1 to 0.01) |
| **Unsure vs. pre-menopausal** | -8.37e-2 (-2.06e-1 to 0.04) |
| **Alcohol consumption** | 7.94e-3 (6.28e-3 to 0.10)* |

**STable 1:** Associations between menopause status and left putamen susceptibility in females with available menopause status at imaging: pre-menopausal (n=580), post-menopausal (n=8129), unclear due to hysterectomy (n=929), unclear for other reasons (n=372). Estimates represent standard deviations and were generated from regression models adjusted for: age, sex, smoking, body mass index, diabetes, cholesterol, blood pressure. *p<0.05.

| **Brain region** | **IVW (95 CI)** |
| --- | --- |
| **Right putamen** | 0.18(0.001 to 0.35)* |
| **Left putamen** | 0.11(-0.05 to 0.27) |
| **Right caudate** | -0.39(-1.00 to 0.21) |
| **Left caudate** | -0.36(-0.95 to 0.23) |
| **Right thalamus** | -0.01(-0.18 to 0.16) |
| **Left thalamus** | 0.01(-0.15 to 0.17) |
| **Right hippocampus** | -0.04 (-0.23 to 0.14) |
| **Left hippocampus** | -0.03(-0.21 to 0.15) |
| **Right substantia nigra** | -0.47(-1.15 to 0.20) |
| **Left substantia nigra** | -0.48(-1.12 to 0.15) |
| **Right amygdala** | -0.07(-0.24 to 0.11) |
| **Left amygdala** | 0.03(0-0.13 to 0.19) |
| **Right pallidum** | -0.49(-1.25 to 0.28) |
| **Left pallidum** | -0.47(-1.23 to 0.29) |
| **Right accumbens** | 0.15(-0.10 to 0.40) |
| **Left accumbens** | 0.12(-0.20 to 0.43) |

**STable 2:** Two-sample Mendelian randomization estimates (two-sample design) for the association of genetically-predicted alcohol use disorder (Psychiatric Genomics Consortium & Million Veterans Program) and quantitative susceptibility mapping image-derived brain phenotypes (UK Biobank). Abbreviations: IVW – inverse-variance weighted, CI – confidence interval. * p value <0.05.

| **Task** | **TMT A** | **TMT B** | **Fluid intelligence** | **Prospective memory** | **Reaction time** | **Grip strength, R** | **Grip strength, L** |
| --- | --- | --- | --- | --- | --- | --- | --- |
| **Putamen, R** | 5.52e-3(3.40e-3 to 7.64e-3)*** | 3.72e-3(1.58e-3 to 5.86e-3)*** | -3.87e-3(-6.03e-3 to -1.72e-3)*** | 3.22e-4(-1.34e-4 to 1.98e-3) | 1.53e-3(-6.36e-4 to 3.70e-3) | -1.14e-3(-2.71e-3 to 4.35e-4) | -1.07e-3(-2.62e-3 to 4.90e-4) |
| **Putamen, L** | 4.71e-3(2.60e-3 to 6.82e-3)*** | 3.82e-3((1.69e-3 to 5.95e-3)*** | -4.08e-3(-6.22e-3 to -1.94e-3)*** | 3.30e-4(-1.32e-3 to 1.98e-3) | 1.27e-3(-8.88e-4 to 3.43e-3) | -6.25e-4(-2.19e-3 to 9.37e-4) | -7.99e-4(-2.35e-3 to 7.51e-4) |
| **Caudate, R** | 2.87e-3(7.66e-4 to 4.97e-3)*** | 3.37e-3(1.25e-3 to 5.49e-3)*** | -2.33e-3(-4.47e-3 to -1.93e-4)** | 3.49e-4(-1.30e-3 to 1.99e-3) | 1.65e-3(-4.94e-4 to 3.80e-3) | -6.18e-5(-1.62e-3 to 1.49e-3) | 1.67e-4(-1.38e-3 to 1.71e-3) |
| **Caudate, L** | 2.77e-3(-2.88e01 to -1.07e-2)** | 2.23e-3(9.85e-5 to 4.37e-3) | -2.03e-3(-4.18e-3 to 1.20e-4) | -7.64e-4(-2.42e-3 to 8.91e-4) | 1.09e-3(-1.26e-3 to 3.06e-3) | -4.94e-4(-1.68e-3 to 1.42e-3) | -1.30e-4(-2.06e-3 to 1.07e-3) |
| **Substantia nigra, R** | 1.34e-4 (-1.93e-3 to 2.20e-3) | 1.98e-3-9.90e-5 to 4.07e-3) | -1.06e-3-3.16e-3 to 1.04e-3) | 7.44e-3(-8.70e-2 to 2.36e-3) | 4.09e-5(-2.07e-3 to 2.15e-3) | 1.05e-3(-4.72e-4 to 2.58e-3) | 1.05e-3(-4.67e-4 to 2.56e-3) |
| **Hippocampus, R** | 3.96e-3(8.43e-4 to 5.05e-3)** | -2.21e-5(-2.15e-3 to 2.11e-3) | -1.60e-3(-3.74e-3 to 5.41e-4) | 4.67-4(-1.18e-3 to 2.12e-3) | 3.77e-4(-1.78e-3 to 2.53e-3) | 1.14e-3(-4.21e-4 to 2.70e-3) | 8.43e-4(-7.04e-4 to 2.39e-3) |
| **Thalamus, R** | 3.68e-4(-1.74e-3 to 2.48e-3) | -3.89e-4(-2.52e-3 to 1.74e-3) | -2.95e-4(-2.44e-3 to 1.85e-3) | 9.02e-4(-7.50e-4 to 2.55e-3) | -1.38e-5(-2.17e-3 to 2.14e-3) | 1.54e-3(6.10e-4 to 3.74e-3)** | 2.17e-3(-1.02e-5 to 3.09e-3) |
| **Thalamus, L** | 1.10e-3  (-1.00e-3 to 3.19e-3) | 1.76e-4  (-1.94e-3 to 2.30e03) | -3.92e-4  (-2.53e-3 to 1.74e-3) | -9.91e-5  (-1.74e-3 to 1.54e-3) | -6.48e-4  (-2.79e-3 to 1.50e-3) | 7.87e-5  (-1.88e-4 to 2.90e-3) | 7.93e-5(1.10e-4 to 3.22e-3) |

**STable 3:** Interactions between age and susceptibility imaging-derived phenotypes in predicting cognitive or motor performance at time of scan. Imaging and cognitive/motor scores were quantile normalized. Estimates represent standard deviation changes and were generated from regression models adjusted for: age, sex, imaging site, education, income, historical job code, Townsend Deprivation Index, smoking, blood pressure, body mass index. Abbreviations: TMT – Trail-making task, R – right, L – left. * p value <0.01, ** <0.001.

| **Task** | **TMT A** | **TMT B** | **Fluid intelligence** | **Prospective memory** | **Reaction time** | **Grip strength, R** | **Grip strength, L** |
| --- | --- | --- | --- | --- | --- | --- | --- |
| **Putamen, R** | -2.16e-3(-1.88e-2 to 1.44e-2) | -6.33e-3(-2.31e2 to 1.04e-2) | 6.92e-3(-9.96e-3 to 2.38e-2) | 3.14e-3(9.86e-3 to 1.61e-2) | 5.62e-3(-1.13e-2 to 2.26e-2) | 1.58e-2(3.53e-3 to 2.81e-2)* | 1.74e-2(5.19e-3 to 2.96e-2)** |
| **Putamen, L** | -9.40e-4(-1.76e-2 to 1.57e-2) | -3.58e-3(-2.04e-2 1.32e-2) | -2.73e-3(-1.97e-2 to 1.42e-2) | 2.26e-3(-1.08e-2 to 1.53e-2) | 1.00e-2(-6.98e-3 to 2.70e-2) | 1.52e-2(2.85e-3 to 2.75e-2) | 1.44e-2(2.17e-3 to 2.66e-2) |
| **Caudate, R** | -1.09e-2(-2.69e-2 to 5.09e-3) | -1.99e-3(-1.81e-2 to 1.41e-2) | -6.16e-3(-2.24e-2 to 1.01e-2) | 1.55e-4(-1.23e-2 to 1.27e-2) | -3.81e-3(-2.01e-2 to 1.25e-2) | 1.55e-2(3.71e-3 to 2.74e-2)* | 1.47e-2(3.00e-3 to 2.65e-2) |
| **Caudate, L** | -8.88e-3(-2.48e-2 to 7.06e-3) | -5.25e-3(-2.13e-2 to 1.09e-2) | -3.50e-3(-1.97e-2 to 1.27e-2) | -6.69e-3(1.92e-2 to 5.79e-3) | -3.80e-3(-2.01e-2 to 1.25e-2) | 1.40e-2(2.18e-3 to 2.58e-2) | 1.07e-2(-1.05e-3 to 2.24e-2) |
| **Substantia nigra, R** | 1.12e-2(-4.39e-3 to 2.68e-2) | 6.26e-3(-9.48e-3 to 2.20e-2) | 1.14e-3(-1.44e-2 to 1.73e-2) | 2.96e-3(-9.23e-3 to 1.52e-2) | 2.36e-2(7.64e-3 to 3.95e-2)** | 8.36e-3(-3.17e-3 to 1.99e-2) | 9.60e-3(-1.85e-3 to 2.10e-2) |
| **Hippocampus, R** | 2.20e-2(5.94e-3 to 3.89e-2)** | 1.81e-2(1.90e-3 to 3.43e-2) | -1.86e-2(-3.49e-2 to -2.30e-3) | 3.90e-3(-8.64e-3 to 1.64e-2) | 1.72e-2(8.01e-4 to 3.36e-2) | -8.58e-3(-2.04e-2 to 3.29e-3) | -8.95e-3(-2.08e-2 to 2.82e-3) |
| **Thalamus, R** | 1.07e-2(-5.22e-3 to 2.67e-2) | 4.70e-3(-1.14e-2 to 2.08e-2) | -1.72e-2(-3.34e-2 to -9.75e-4) | 6.62e-4(-1.18e-2 to 1.32e-2) | 1.60e-2(-3.05e-4 to 3.23e-2) | 9.21e-3(-2.61e-3 to 2.10e-2) | 5.58e-3(-6.15e-3 to 1.73e-2) |
| **Thalamus, L** | 8.84e-3(-7.09e-3 to 2.48e-2) | 1.51e-3-1.46e-2 to 1.76e-2) | -7.23e-3(-2.34e-2 to 8.96e-3) | -5.59e-3(-1.81e-2 to 6.87e-3) | 1.31e-2(-3.17e-3 to 2.94e-2) | 4.78e-3(-7.01e-3 to 1.66e-2) | -7.21e-4-1.24e-2 to 1.10e-2) |

**STable 4:** Associations between brain susceptibility image-derived phenotypes and cognitive and motor tests at time of scan. Imaging and cognitive/motor scores were quantile normalized. Estimates represent standard deviation changes and were generated from regression models adjusted for: age, sex, imaging site, education, income, historical job code, Townsend Deprivation Index, smoking, blood pressure, body mass index. Abbreviations: TMT – Trail-making task, R – right, L – left. * p value <0.01, ** <0.001.
