## Supplementary figures for "Impact of moderate alcohol consumption on brain iron and cognition: observational and genetic analyses"

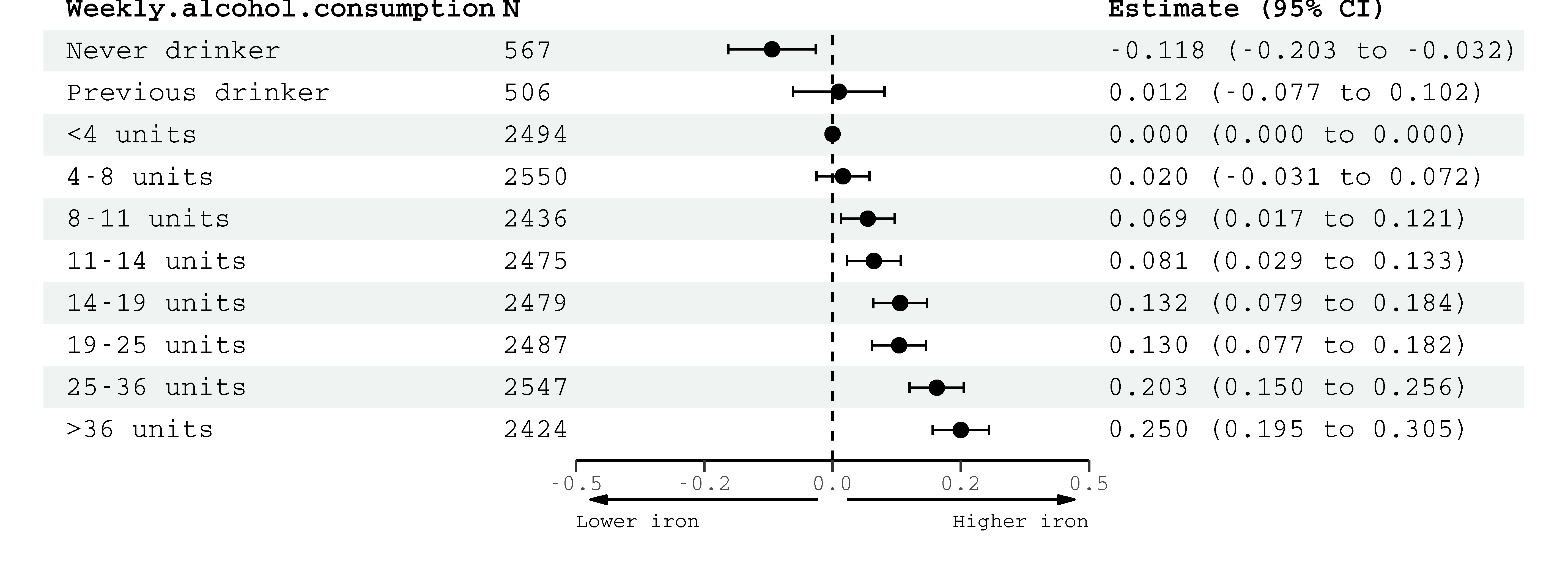

**Figure 1: Left putamen susceptibility** associations with alcohol consumption (octiles, reference group = <4 units weekly). Estimates (95% confidence intervals) represent standard deviations and were generated from regression models adjusted for: age, sex, smoking, body mass index, diabetes, cholesterol, blood pressure and imaging-related confounds.

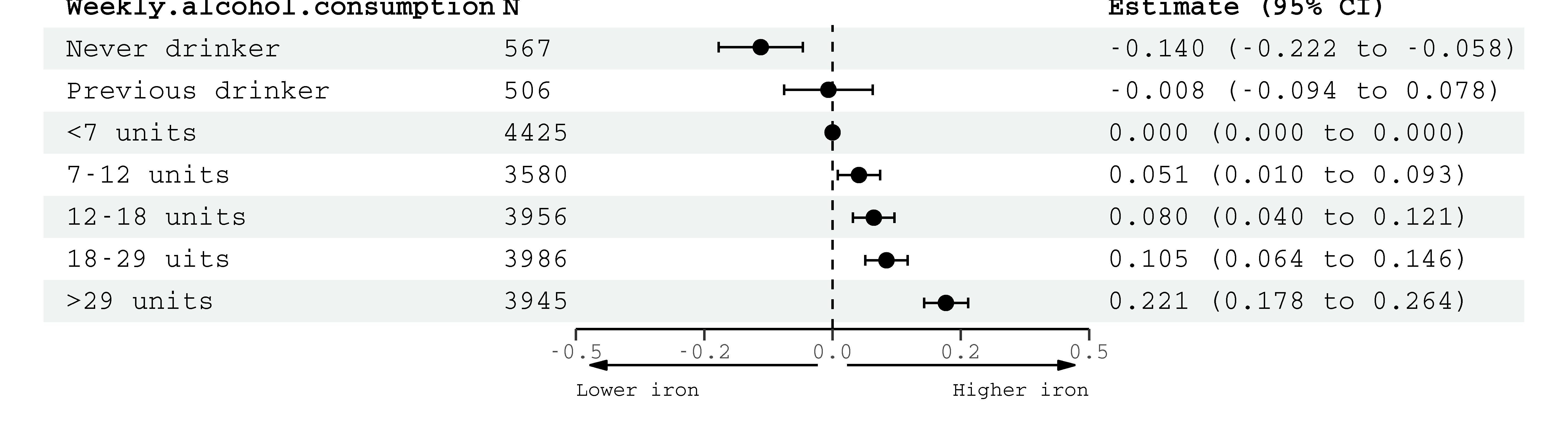

**Figure 2: Right putamen susceptibility** associations with alcohol consumption (quintiles, reference group = <7 units weekly). Estimates (95% confidence intervals) represent standard deviations and were generated from regression models adjusted for: age, sex, smoking, body mass index, diabetes, cholesterol, blood pressure and imaging-related confounds.

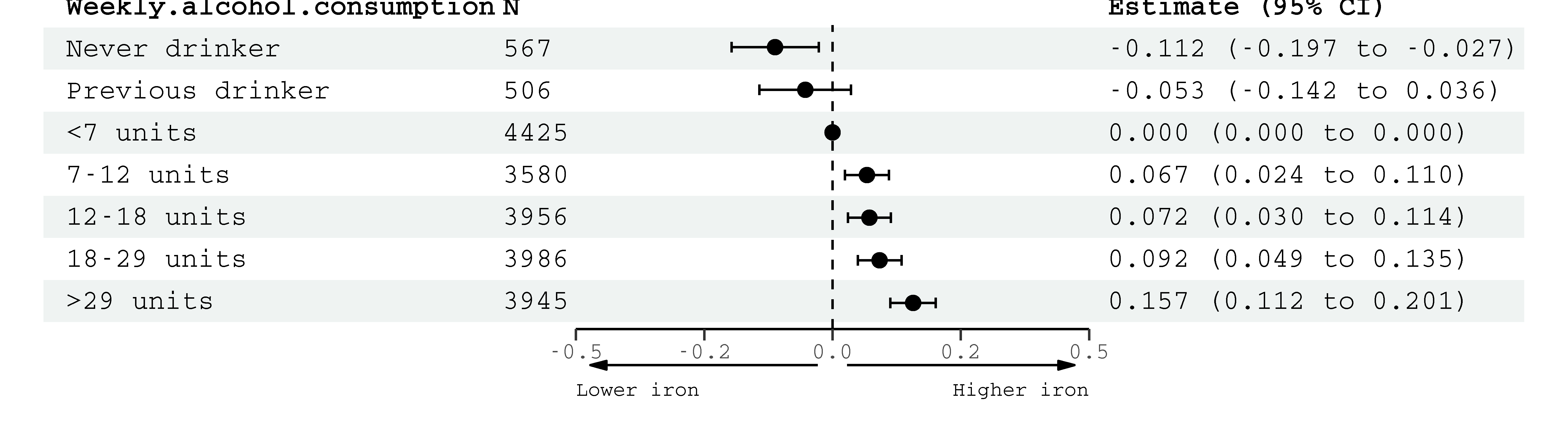
**Figure 3: Right caudate susceptibility** associations with alcohol consumption (quintiles, reference group = <7 units weekly). Estimates (95% confidence intervals) represent standard deviations and were generated from regression models adjusted for: age, sex, smoking, body mass index, diabetes, cholesterol, blood pressure and imaging-related confounds.

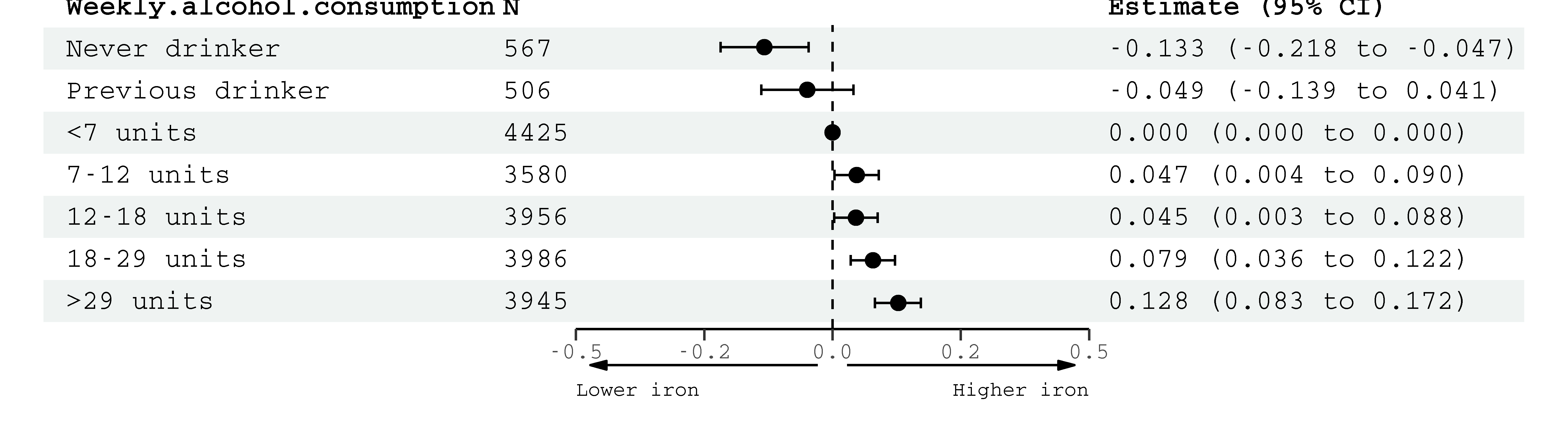

**Figure 3: Left caudate susceptibility** associations with alcohol consumption (quintiles, reference group = <7 units weekly). Estimates (95% confidence intervals) represent standard deviations and were generated from regression models adjusted for: age, sex, smoking, body mass index, diabetes, cholesterol, blood pressure and imaging-related confounds.

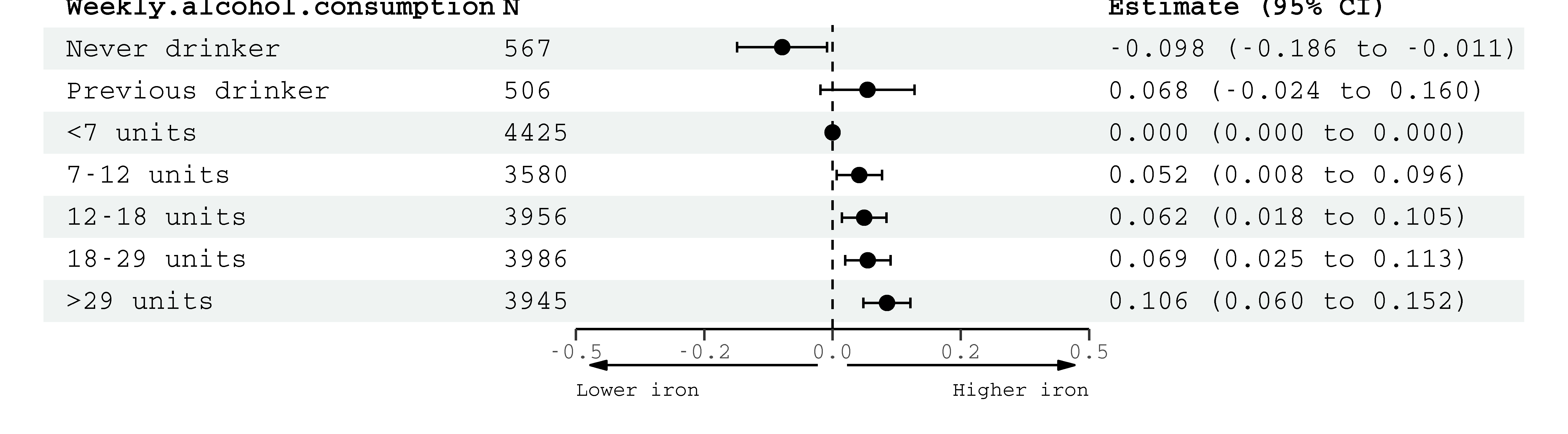

**Figure 4: Left substantia nigra susceptibility** associations with alcohol consumption (quintiles, reference group = <7 units weekly). Estimates (95% confidence intervals) represent standard deviations and were generated from regression models adjusted for: age, sex, smoking, body mass index, diabetes, cholesterol, blood pressure and imaging-related confounds.

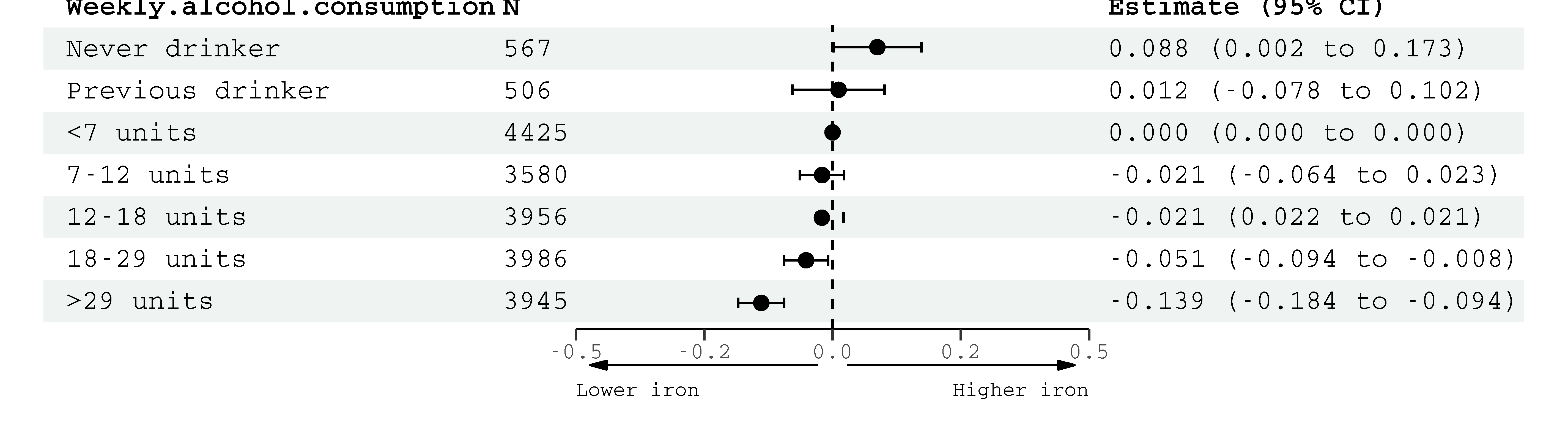

**Figure 5: Right thalamus susceptibility** associations with alcohol consumption (quintiles, reference group = <7 units weekly). Estimates (95% confidence intervals) represent standard deviations and were generated from regression models adjusted for: age, sex, smoking, body mass index, diabetes, cholesterol, blood pressure and imaging-related confounds

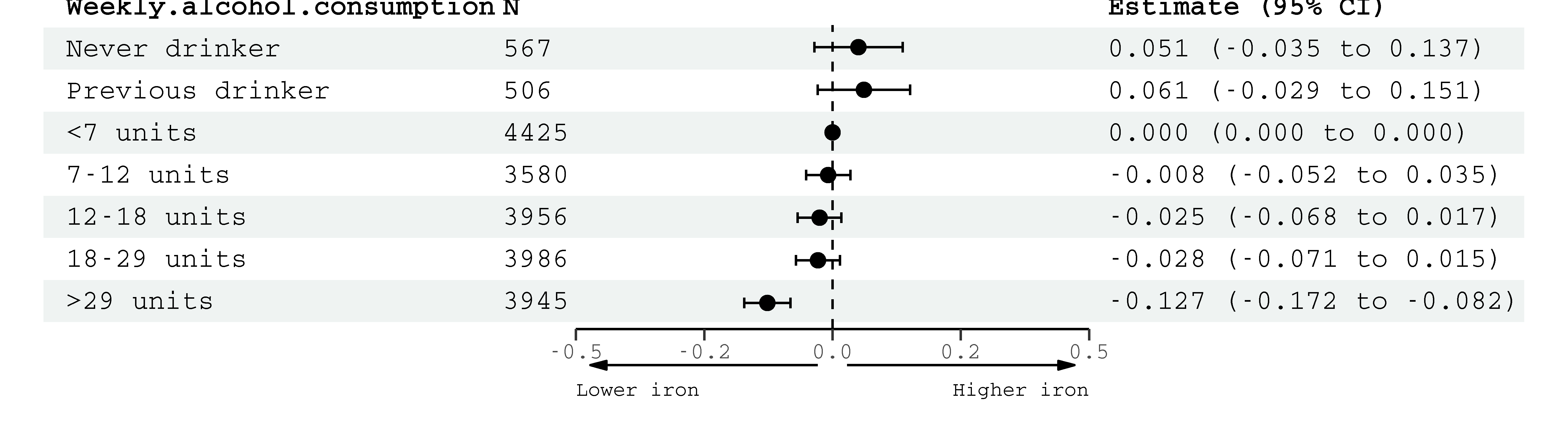

**Figure 6: Left thalamus susceptibility** associations with alcohol consumption (quintiles, reference group = <7 units weekly). Estimates (95% confidence intervals) represent standard deviations and were generated from regression models adjusted for: age, sex, smoking, body mass index, diabetes, cholesterol, blood pressure and imaging-related confounds.

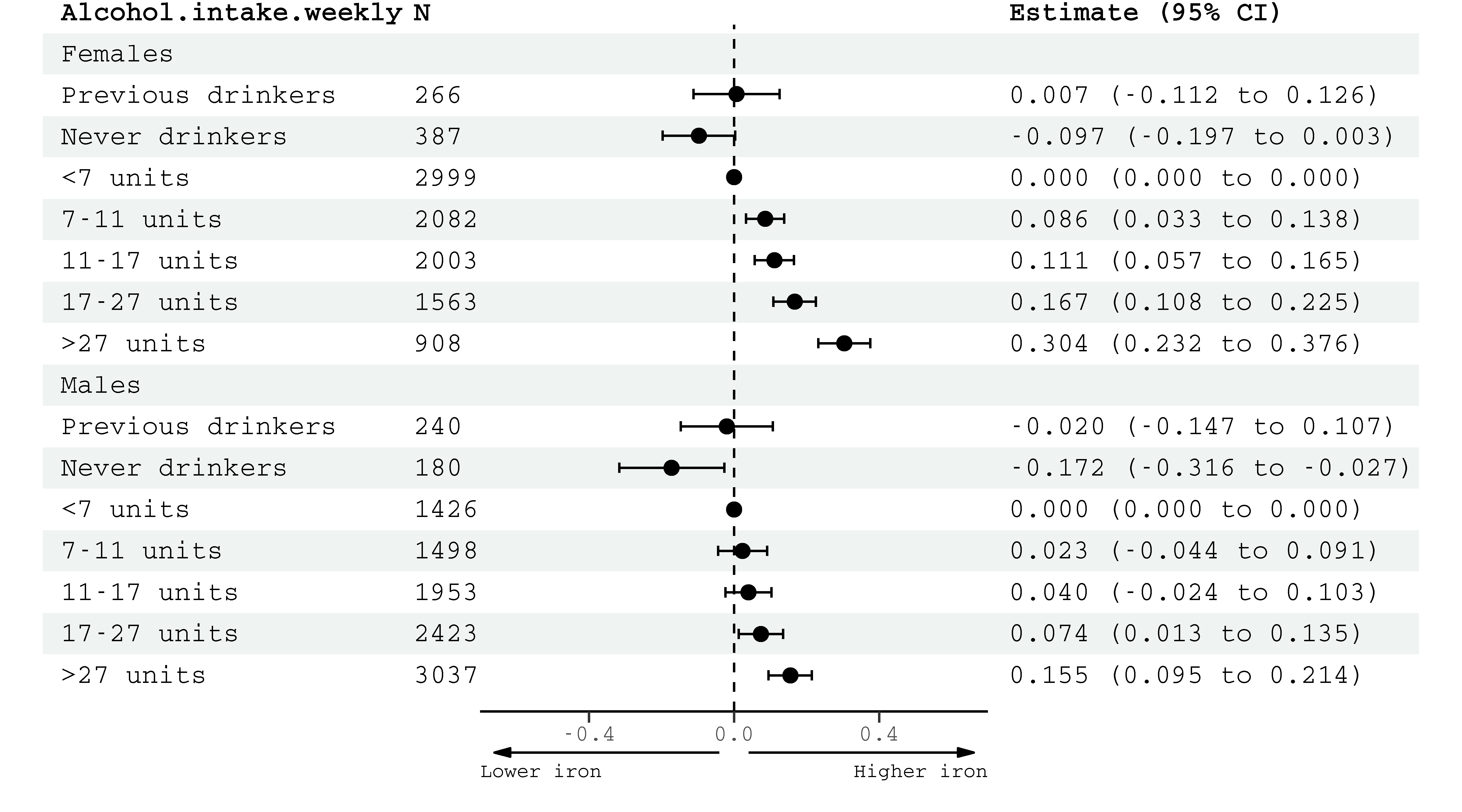

**Figure 7: Left putamen susceptibility** associations with alcohol consumption (quintiles, reference group = <7 units weekly) by sex. Estimates (95% confidence intervals) represent standard deviations and were generated from regression models adjusted for: age, sex, smoking, body mass index, diabetes, cholesterol, blood pressure and imaging-related confounds.

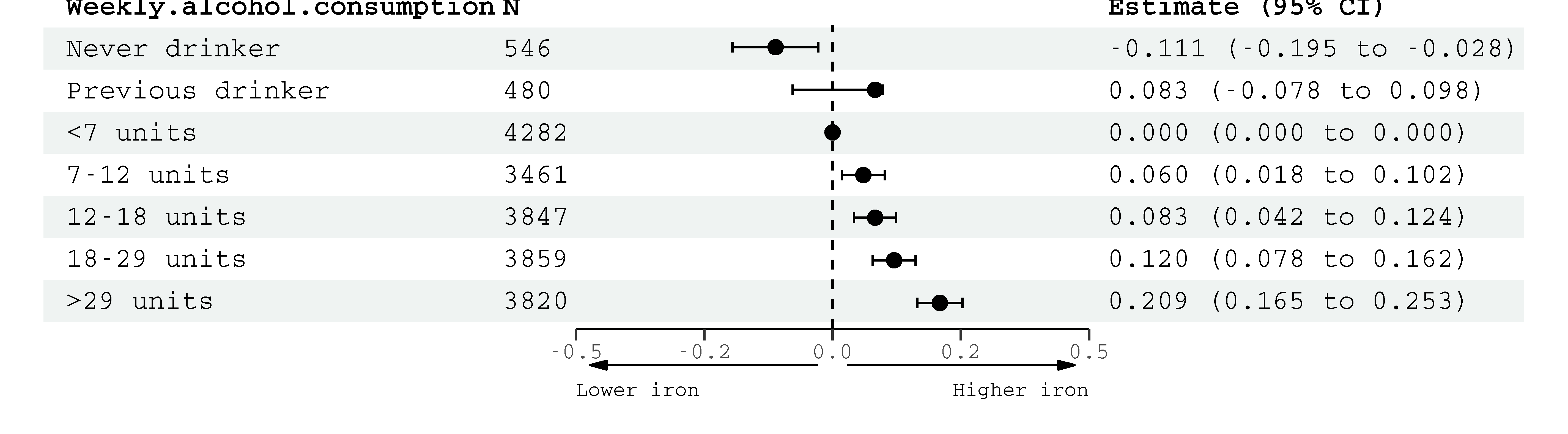

**Figure 8: Left putamen susceptibility** associations with alcohol consumption (quintiles, reference group = <7 units weekly). Estimates (95% confidence intervals) represent standard deviations and were generated from regression models adjusted for: age, sex, smoking, body mass index, diabetes, cholesterol, blood pressure and imaging-related confounds and additional adjustment for diet – red meat, fish, vegetables, fruit and bread consumption frequency weekly at time of the scan.

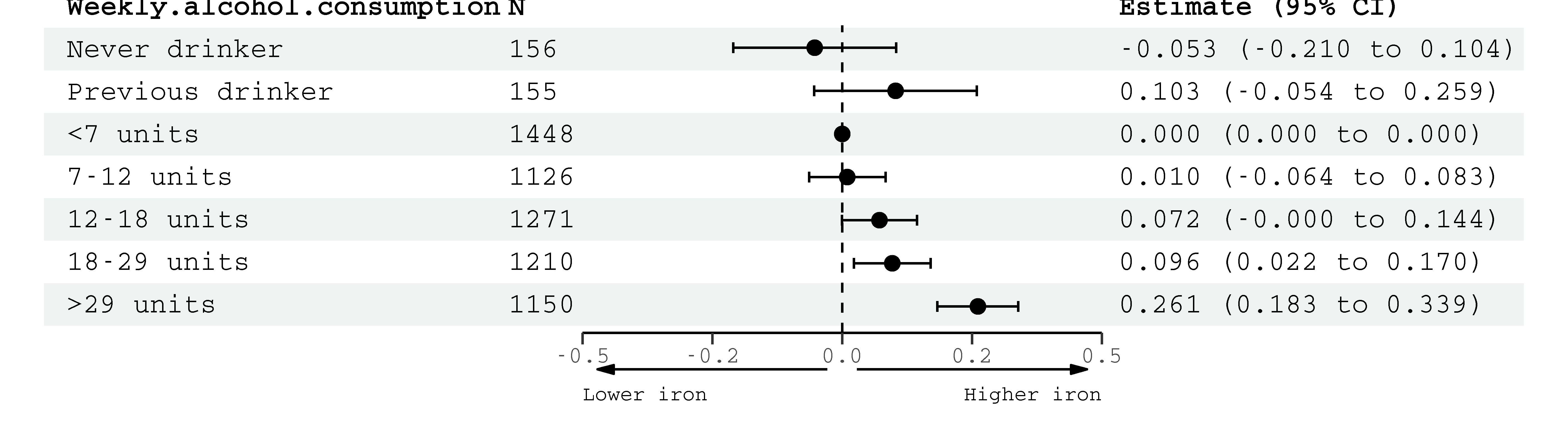

**Figure 9: Left putamen susceptibility** associations with alcohol consumption (quintiles, reference group = <7 units weekly). Estimates (95% confidence intervals) represent standard deviations and were generated from regression models adjusted for: age, sex, smoking, body mass index, diabetes, cholesterol, blood pressure and imaging-related confounds and additional adjustment for diet (red meat, fish, vegetables, fruit and bread consumption frequency weekly) and iron supplementation at time of the scan.

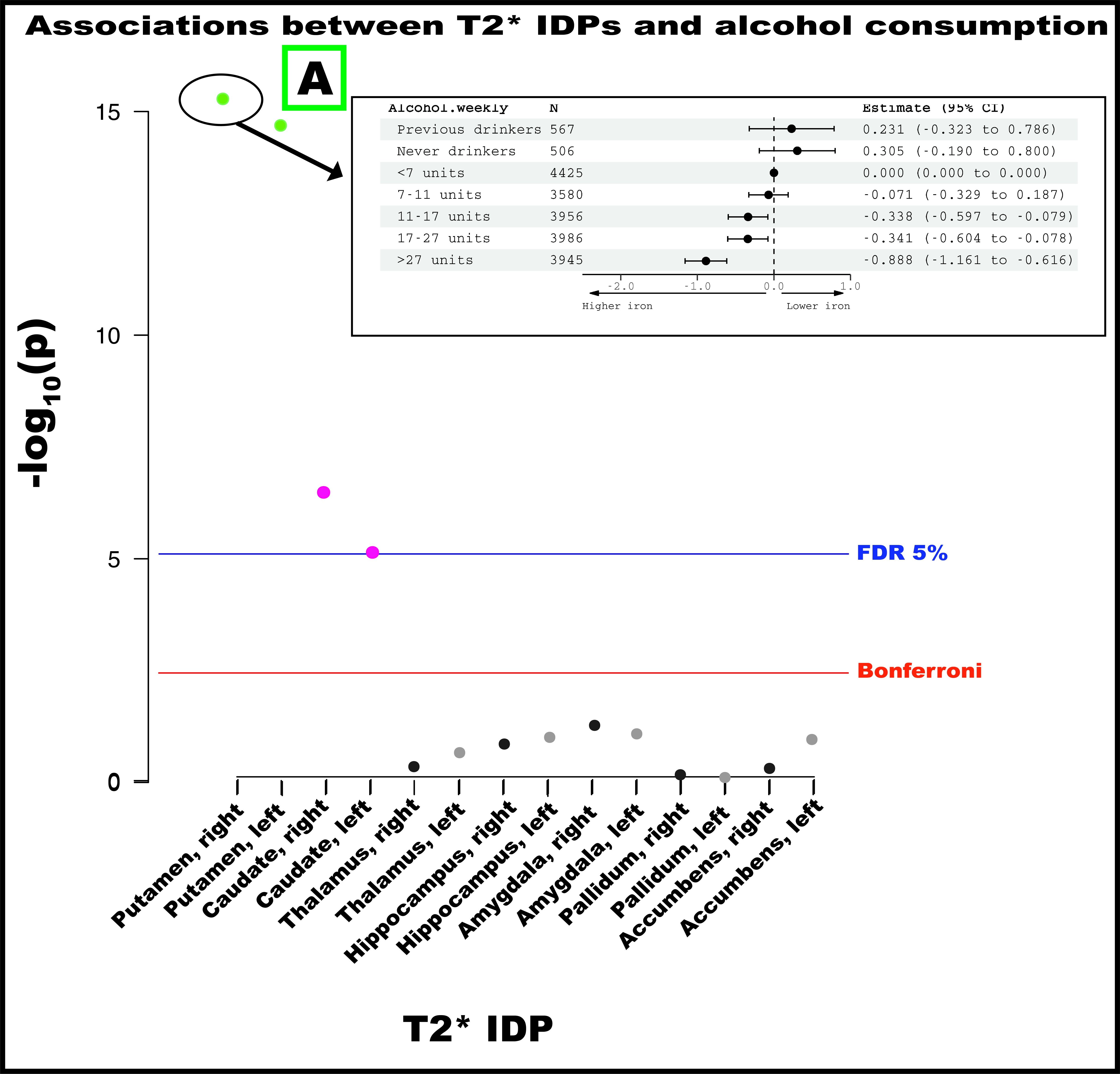

**SFigure 10:** Associations between weekly alcohol intake and T2* imaging-derived phenotypes. Insert **(A)** shows associations between alcohol intake (sample quintiles) and right putamen T2* where the reference group are those drinking <7 units (56g) weekly (reference, estimates (95% CI) set to 0). Estimates represent standard deviations and were generated from regression models adjusted for: age, sex, smoking, body mass index, diabetes, cholesterol, blood pressure, rs1800562, rs1799945 and rs855791 and full set of imaging-related confounds. Blue line indicates False Discovery Rate threshold (5%) and red line Bonferroni threshold. Abbreviations: IDP – imaging-derived phenotype, FDR – false discovery rate.

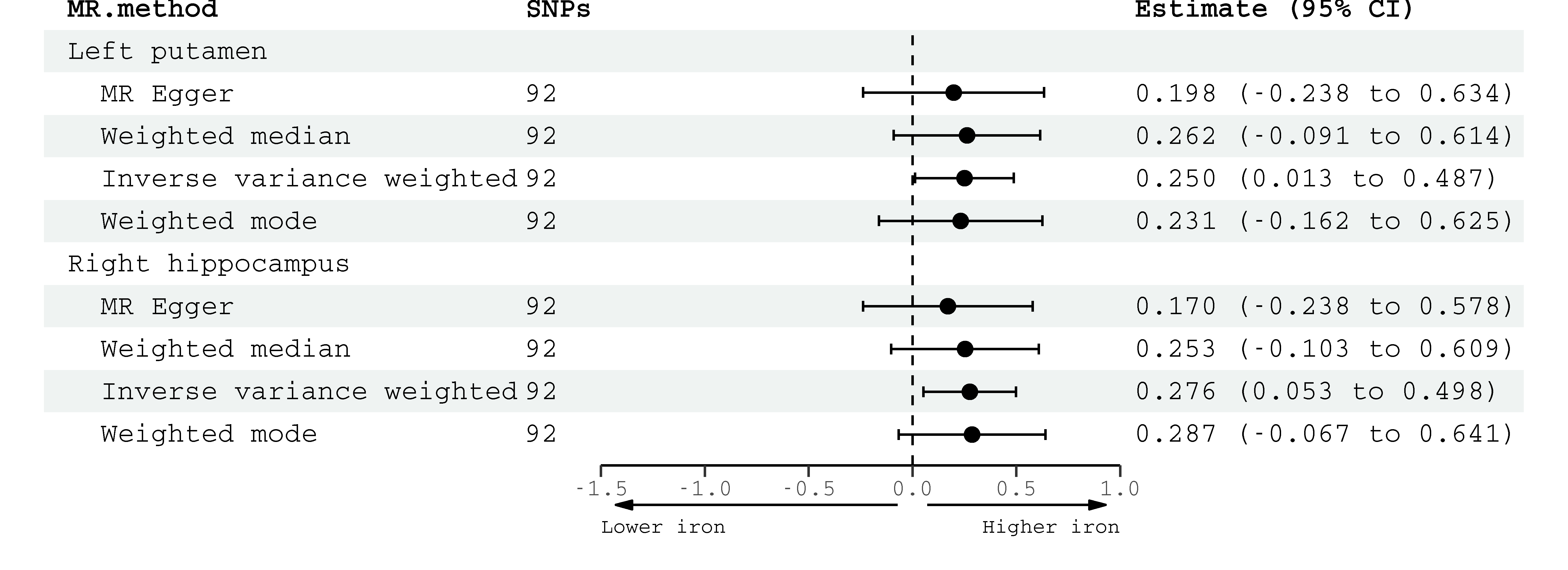

**SFigure 11:** Robust Mendelian Randomization estimates (two-sample design) for significant associations between genetically predicted alcohol consumption (GWAS & Sequencing Consortium of Alcohol and Nicotine) and susceptibility imaging-derived phenotypes (UK Biobank) in inverse-variance weighted analysis (main **Figure 3**). Effect estimates for alcohol consumption are per standard deviation increase in genetically-predicted log-transformed drinks per week. Abbreviations: MR – Mendelian randomization, SNPs – single nucleotide polymorphisms, CI – confidence interval.

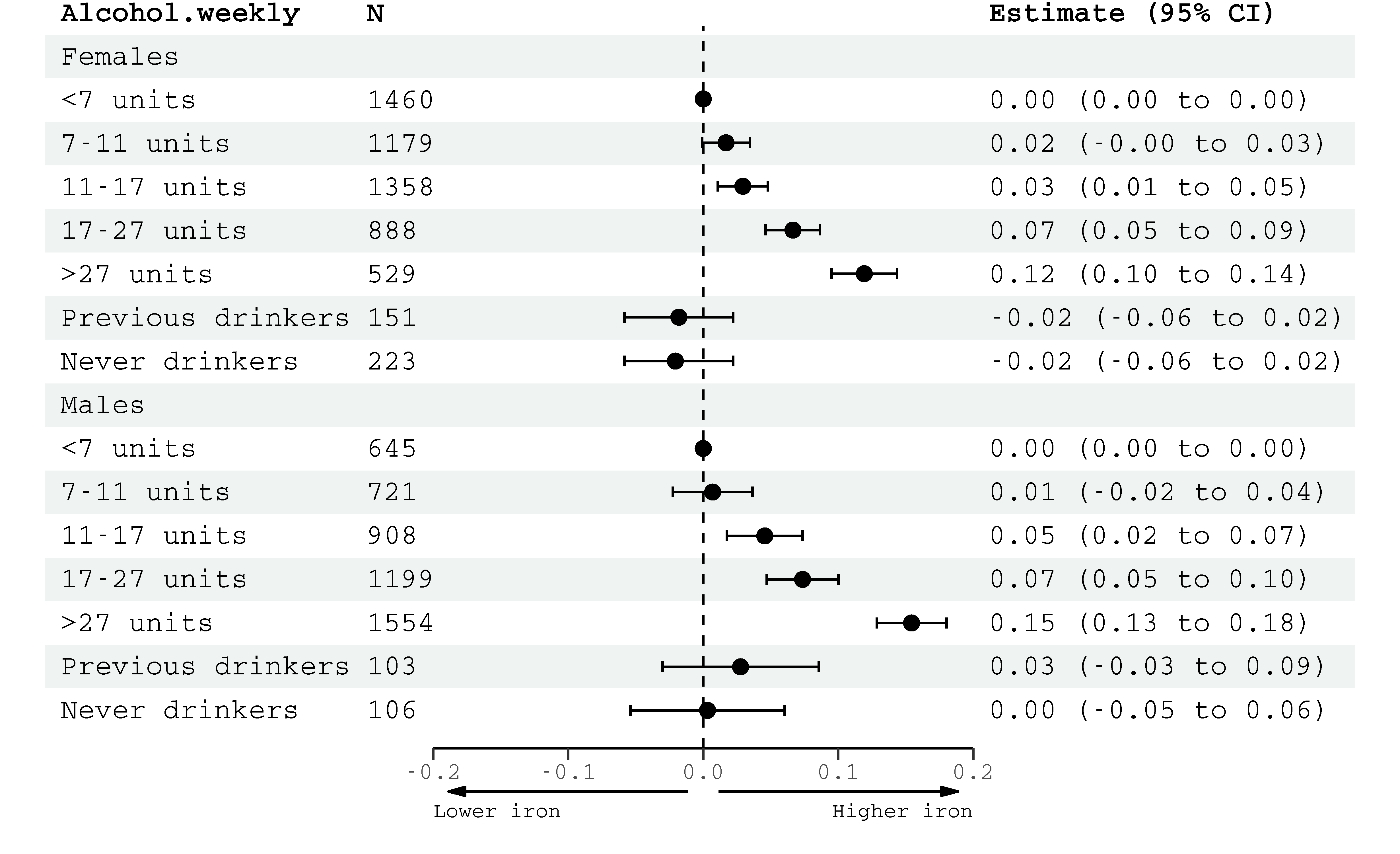

**SFigure 12:** Associations between weekly alcohol consumption (quintiles) and liver iron (mg/g). Reference group is those drinking <7 units (56g) weekly. Estimates generated from regression models (n=11,024) adjusted for: age, smoking, imaging site, diabetes mellitus, body mass index, blood pressure, cholesterol, dietary iron, rs1800562, rs1799945 and rs855791. N=11,024.

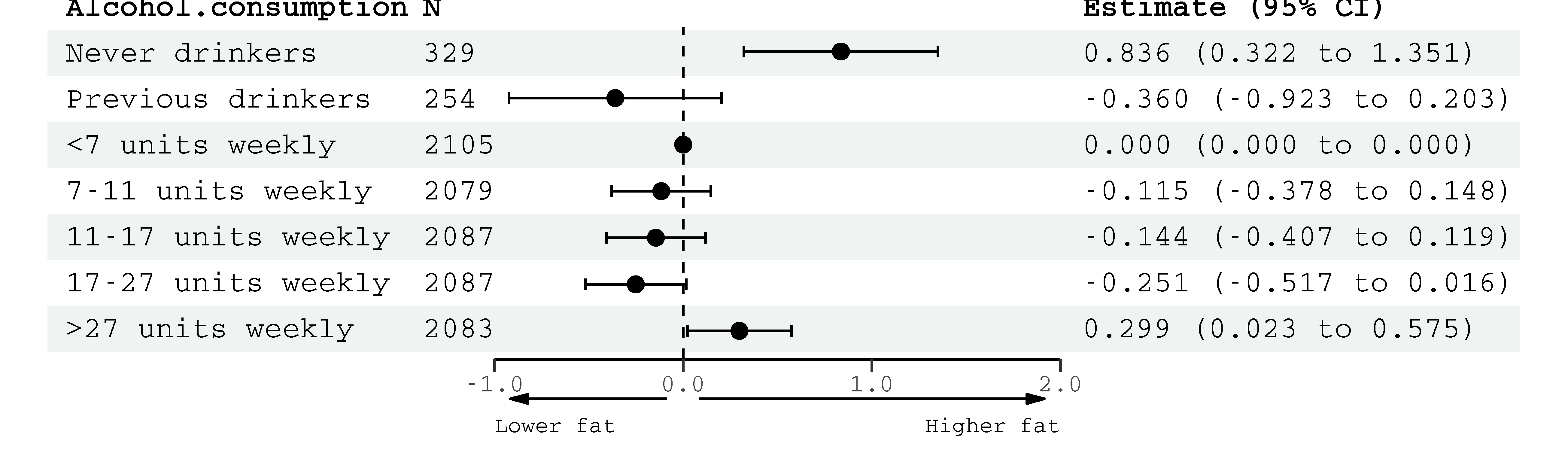

**SFigure 13:** Associations between **a**lcohol consumption weekly (quintiles) and liver protein density fat fraction (PDFF, %). Estimates are adjusted for age, sex, smoking, body mass index, cholesterol, blood pressure, education, Townsend Deprivation Index, household income, historical job type.

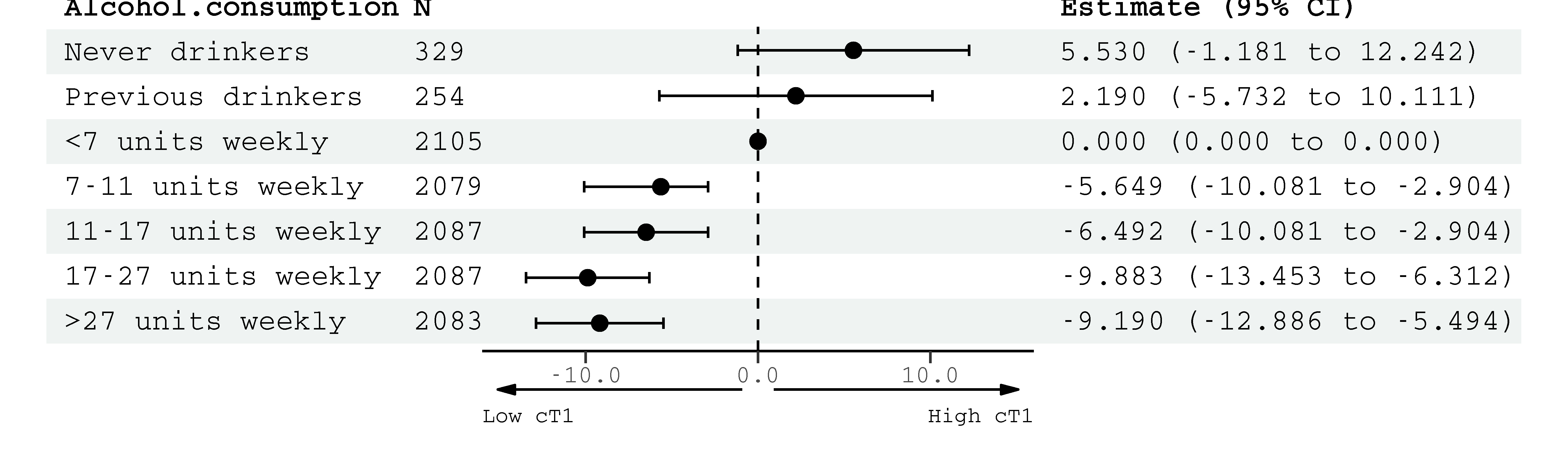

**SFigure 14:** Associations between alcohol consumption weekly (quintiles) and liver cT1 (milliseconds), a marker of inflammation/fibrosis. Estimates are adjusted for: age, sex, smoking, body mass index, cholesterol, blood pressure, education, Townsend Deprivation Index, household income, historical job type. All estimates are within the normal reference range [1].

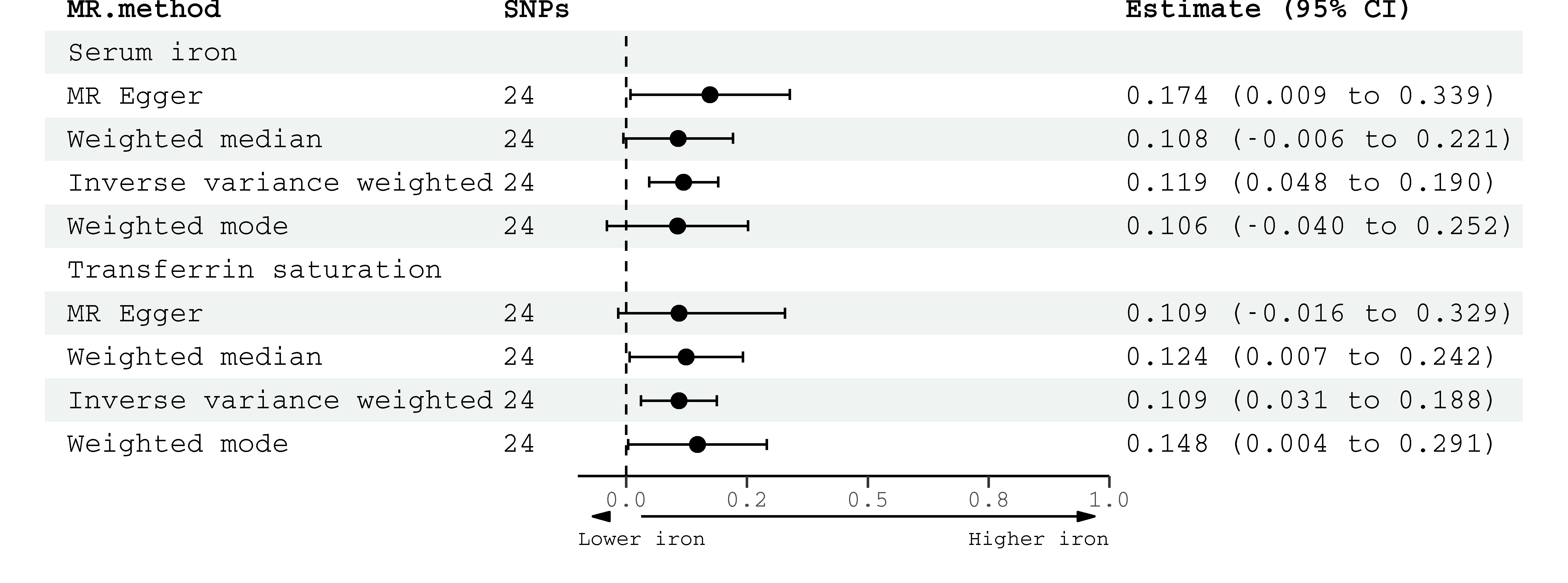

**SFigure 15:** Robust Mendelian randomization estimates (two-sample design) for the association of genetically predicted alcohol use disorder (Million Veterans Program & Psychiatric Genomics Consortium) and serum markers of iron homeostasis (deCODE, INTERVAL and Danish Blood Donor Study).

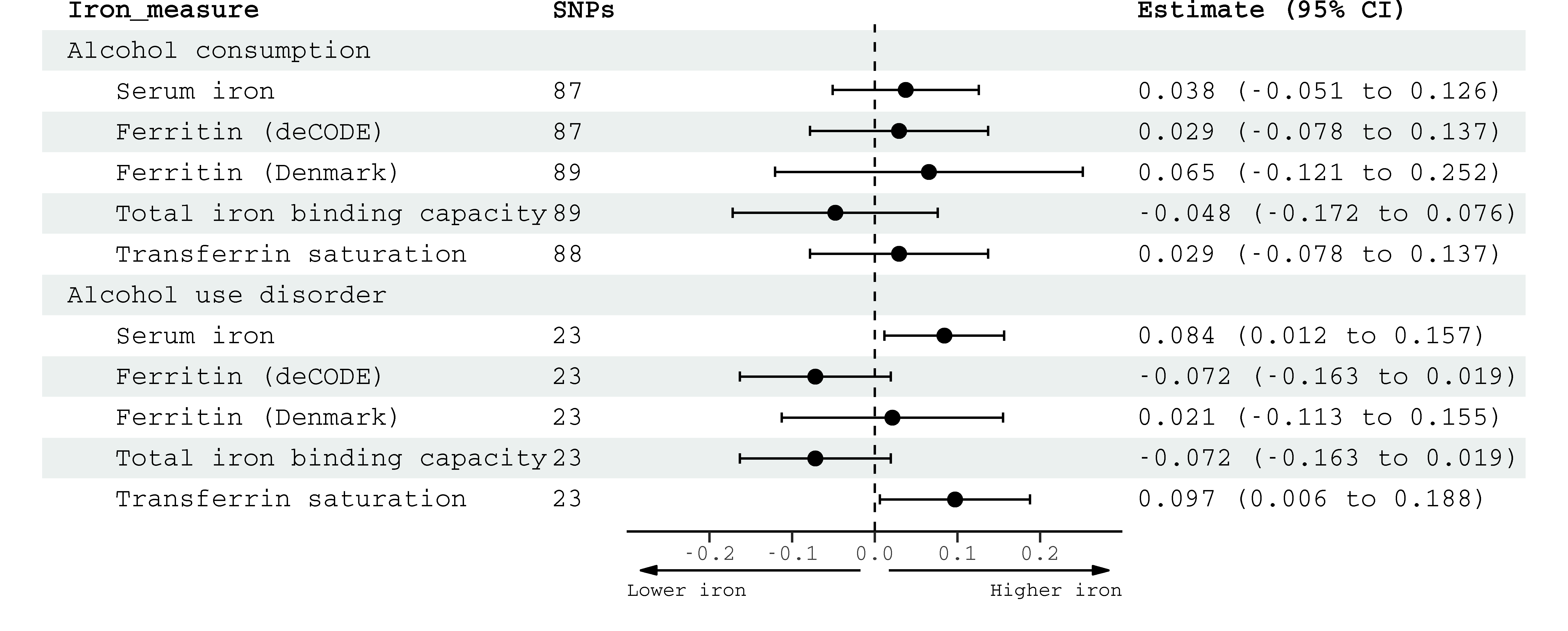

**SFigure 16:** Mendelian randomization (two-sample design) estimates for the associations of genetically predicted alcohol consumption (log-transformed drinks per week, GWAS & Sequencing Consortium of Alcohol and Nicotine)/Alcohol Use Disorder (Million Veterans Program & Psychiatric Genomics Consortium) with serum iron markers, in cohorts that did not adjust for alcohol in their genome-wide association study (DECODE unless otherwise marked).

1.	Mojtahed A, Kelly C, Herlihy A, Kin S, Wilman H, Mckay A, et al. Reference range of liver corrected T1 values in a population at low risk for fatty liver disease—a UK Biobank sub-study, with an appendix of interesting cases. Abdominal Radiology. 2019;44(1):72-84.
